# A TALS-like *RTTN* mutation impedes neural rosette formation in human cortical organoids

**DOI:** 10.1101/2024.04.03.24303866

**Authors:** Justine Guguin, Ting-Yu Chen, Alicia Besson, Silvestre Cuinat, Eloïse Bertiaux, Lucile Boutaud, Nolan Ardito, Miren Imaz Murguiondo, Sara Cabet, Virginie Hamel, Sophie Thomas, Bertrand Pain, Patrick Edery, Audrey Putoux, Tang K. Tang, Sylvie Mazoyer, Marion Delous

## Abstract

The Taybi-Linder syndrome (TALS) is a rare genetic disorder characterized by a severe microcephaly with abnormal gyral pattern, severe growth retardation, bone abnormalities and a reduced life span for the most severe cases. It is caused by mutations in *RNU4ATAC* whose transcript, the small nuclear RNA U4atac, is a core component of the minor spliceosome involved in the excision of minor introns spread over ∼750 genes.

Here, we report a patient presenting with TALS features but no mutation in *RNU4ATAC*; instead, she carries the *RTTN* c.2953A>G variant at the homozygous state. This variant, already reported in patients with syndromic microcephaly, encodes the missense p.Arg985Gly amino acid change. It is also known to affect *RTTN* pre-mRNA splicing, with the expression of two forms lacking either exon 23 (in-frame) or exons 22-23 (out-of-frame). By using the engineered *RTTN* depleted RPE1 cellular model, we analysed independently the impact of the missense and in-frame deletion of exon 23 *RTTN* isoforms on the localisation and function of the protein at the centrosome, and showed that the pathogenicity of the c.2953A>G variant is mostly due to the latter. In patient fibroblasts, we observed a reduction of the centriole length and an alteration of ciliary function, while the analysis of neuronal stem cells (NSC) derived from CRISPR/Cas9-edited induced pluripotent stem cells revealed major cell cycle and mitotic abnormalities, leading to aneuploidy, cell cycle arrest and increased cell death. Finally, by generating cortical organoids, we discovered a new function of RTTN in the self-organisation of NSC into neural rosettes. We observed a delayed apico-basal polarization of NSC, accompanied with decreased cell division and increased apoptosis. Altogether, these defects lead to a marked decrease of rosette number and size in *RTTN*-mutated organoids, thus impeding their overall growth.

To conclude, our study gives new insights on microcephaly-related pathophysiological mechanisms underlying the only recurrent *RTTN* mutation, that could also open a path to better understand those involved in *RNU4ATAC*-associated Taybi-Linder syndrome.

## Introduction

The Taybi-Linder syndrome (TALS), or MOPD1 (OMIM #210710), is a very rare genetic disease with less than 100 reported cases^1–6^ characterized by severe pre- and post-natal growth delay, severe microcephaly with brain abnormalities (abnormal gyral pattern, intracranial cyst, cerebellar vermis hypoplasia and corpus callosum agenesis), intellectual deficiency, bone abnormalities, bulging eyes and dry skin with eczema, as well as a premature death during the first months of life for the most severe cases.^7–9^ Since the identification in 2011 of *RNU4ATAC*,^10,11^ no other gene has been associated to this disease. *RNU4ATAC* is a non-coding gene transcribed into the small nuclear RNA (snRNA) U4atac, a component of the minor spliceosome. This ribonucleic complex excises less than one percent of all human introns,^12^ and deficiency in its function leads in most cases to minor intron retention that impairs the correct maturation of about 750 minor-intron containing transcripts.^2,13–17^

Here, we report a particular case of a patient with TALS traits but no variant in *RNU4ATAC*. Whole exome sequencing rather revealed she carries a homozygous pathogenic variant in *RTTN*. This gene has already been implicated in human disease, notably “microcephaly, short stature and polymicrogyria with or without seizure” (MSSP, OMIM #614833).^18^ Up to now, 37 patients carrying bi-allelic pathogenic variants in *RTTN* have been reported.^18–32^ All these patients exhibit common features, at variable degrees of severity, including microcephaly with gyral pattern abnormalities, slopping forehead, intellectual deficiency, and pre- and post-natal growth delay. A few patients only were deceased at the time of publication, with one third of them having reached teenage- or adult-hood.

*RTTN* encodes the large centrosomal protein Rotatin (2,226 amino-acids), and was initially discovered by a forward genetic screen in the mouse model, with the particular phenotype of *Rttn* mutant embryos failing to undergo the axial rotation during embryogenesis.^33^ Further investigations demonstrated that Rotatin is essential for procentriole elongation,^34^ correct cell cycle progression and mitosis^30,35^ and regulation of primary cilium length.^19,30^ Alterations of all these processes have been linked to microcephaly.^36,37^

Intrigued by the TALS-like phenotype of the patient with this *RTTN* variant, we thought to investigate the associated pathophysiological mechanisms leading to microcephaly. For that, we used patient fibroblasts, engineered RPE1 cells and CRISPR/Cas9-edited induced pluripotent stem cells (iPSC) that were differentiated into neural stem cells (NSC) or cortical organoids (CO). We carried out a thorough analysis of the impact of the *RTTN* isoforms resulting from the mutated allele identified in the patient, and conducted cellular studies of the centrosome/primary cilium complex and their related functions. Overall, our results establish how *RTTN* c.2953A>G variant affects the integrity of the neuronal stem cell pool, thus leading to improper neuronal cell mass in patients.

## Materials and methods

### Cell culture

Human primary fibroblasts were obtained from the CBC Biotec of the Hospices Civils de Lyon (certified with a specific French standard for biobanks, NF S96-900). Informed written consent for the use of these samples in research was obtained from parents of the *RTTN* case and from those of the sex- and age-matched control. The study on patient cells was approved by the French national ethical committee Comité de Protection des Personnes (number 2021-A01551-40). They were cultivated in HAMF10 medium (Eurobio, CM1H1000-01), complemented with 12% fetal bovine serum (FBS, Eurobio, CVFSVF00-01) and 1% penicillin-streptomycin (PS, Gibco, 15140122). Human telomerase-immortalized retinal pigment epithelial cells (hTERT-RPE1) and derivatives, such as *RTTN^+/+^; p53^+/+^* (control RPE1), *RTTN^+/+^; p53^−/−^* (*p53*-KO), and *RTTN^−/−^; p53^−/−^* (*RTTN*-dKO)-based RTTN-GFP-inducible cells (WT or mutations) (Supplementary Methods) were grown in Dulbecco’s modified Eagle DMEM/F12 medium, supplemented with 10% FBS. For inhibition of nonsense-mediated mRNA decay pathway, fibroblasts were treated with 100 µg/mL cycloheximide for 6 hours. For analysis of primary cilium, fibroblasts and RPE1-derived cell lines were maintained in culture medium with only 0.5% FBS for 48 hours; for cilium disassembly, FBS was reintroduced after 48-hour starvation for the time indicated in the figures. Induced Pluripotent Stem Cells (iPSC) issued from a healthy European male (PCi-CAU2) (Phenocell, Grasse, France) and its CRISPR/Cas9-edited derivatives (Supplementary Methods) were cultivated in 35-mm vitronectin-coated dish (STEMCELL Technologies, 7180) with mTeSR^TM^ Plus medium (STEMCELL Technologies, 100-0276) supplemented with 0,1% PS. Cells were thawed in mTeSR^TM^ Plus medium supplemented with 10 µM ROCK inhibitor Y-27632 (STEMCELL Technologies, 72302).

### iPSC-derived neuronal stem cell and neuron differentiation

The differentiation protocol was adapted from Boutaud *et al.*^38^ with the following adjustments (Supplementary Fig. 7A). After 12 to 14 days in neural induction medium with medium replacement every other day, dense regions appeared and were mechanically detached into small, squared clumps and transferred into a non-coated 35-mm dish with neural expansion medium (NEM) to form neurospheres overnight (Supplementary Table 2). The following day, neurospheres were plated onto Geltrex (Fisher Sci., 15180617) or poly-L-ornithine/laminin (PO/L)-coated dish (Sigma Aldrich, P3655/L2020) and medium changed every other day until neural rosettes appeared. Rosettes were then precisely excised and slightly crumbled before plating onto a Geltrex or PO/L-coated dish (passage 0). From then on, neural stem cells (NSC) were cultivated in NEM and passed at high density (100,000 cells/cm²) every 4 to 5 days using EDTA (Sigma Aldrich, E8008). From passage 3, cells were frozen in cryostore (STEMCELL Technologies, 07930) or cultivated until passage 10 maximum. To generate neurons, NSC were passed at low density (50,000 cells/cm²) using EDTA and cultivated in Geltrex or PO/L-coated dish in N2B27 medium. Medium was changed every other day, and after 7 days in N2B27, 1 µL/mL laminin was added (Sigma Aldrich, L2020). NSC correct differentiation was evaluated through morphological changes (Supplementary Fig. 7A), downregulation of pluripotency genes and upregulation of neuroectodermal genes (Supplementary Fig. 7B) and expression of NSC markers by immunostaining (Supplementary Fig. 7C). For mitotic angles, NSC were blocked with 9 µM RO-3306 (Sigma, SML0569) for 18 hours. Upon release, 5 µM MG-132 (Sigma, M7449) was added to the media for 1 hour before fixation.

### Cortical organoid differentiation and growth curve

iPSC were cultivated in mTeSR+ on vitronectin-coated 35mm dishes until they reached 70% confluence. Using TrypLE Select (Fisher Sci., 11588846), iPSC were dissociated and 2,000 cells per well were plated in u-bottom ultra-low binding 96-well plates (ThermoFisher, 174925) with 150 µL of embryoid body medium (Supplementary Table 3). After seven days, medium was changed toward neural induction medium. At DIV21, organoids were put into differentiation medium containing B27 without vitamin A. At DIV31, B-27 was supplemented with vitamin A (Gibco, 11530536) (Supplementary Fig. 9A). Organoids were harvested at DIV35 for RNA extraction, and at DIV35, DIV46, DIV56 and DIV67 for cryosections and immunostaining. Growth curved was obtained by imaging twenty individual organoids, twice a week, and by measuring their area using Fiji software. Correct differentiation of CO was assessed by RT-qPCR analysis of gene expression of pluripotency and neuroectodermal markers at DIV35 (Supplementary Fig. 9B), and by immunostaining of neuroectodermal markers (PAX6, SOX2, Nestin) at DIV46 (Supplementary Fig. 9C).

### Flow cytometry analysis of cell cycle

Cells were plated at low confluence (20,000 to 50,000 cells/cm²) and harvested with accutase two days later. After cells were washed with PBS, they were fixed for 1h at 4°C in 70% cold ethanol added drop by drop on a vortex. Cells were washed twice with PBS, and stained at room temperature for 30 minutes with 50 µg/mL propidium iodide (Sigma Aldrich, P4170) in presence of 100 µg/mL RNAse A (Sigma Aldrich, R6513). Flow cytometry analysis on 10,000 cells was performed using Canto II and results were analyzed with the FlowJo software v.10.9.0 (BD Biosciences).

### Immunofluorescence staining

Fibroblasts, RPE1 and NSC were respectively seeded on non-coated glass, Geltrex or PO/L-coated coverslips in 24-well plates at low density for cell cycle experiments, or at high density for cilium experiments in HAMF10, DMEME/F12 or neural expansion medium, respectively. When the desired confluence was obtained, medium was removed, and after a wash with PBS, cells were fixed in PBS-4% paraformaldehyde (PFA, EMS, 15713) for 20 minutes at room temperature (fibroblasts, NSC) or with ice-cold methanol at -20 °C for 6 minutes (RPE1). Regarding organoids, they were washed in PBS and fixed in 4% PFA for 4 hours at room temperature with agitation. After two consecutive nights in 15% and in 30% sucrose, they were mounted in cryomolds with OCT embedding matrix and rapidly frozen with liquid nitrogen before being stored at -80°C. Cryosections of 14 µm-width were performed at -30°C with a Micron NX50 cryostat, and sections were conserved at -20°C on superfrost slides. Sections were allowed to defrost 30 minutes at room temperature before being washed three times 10 minutes in PBS-0.5% Triton. Saturation was performed for 1 hour at room temperature with either, for monolayer cells, solution 1 (1X PBS, 10% normal goat serum (Merck Millipore, S26), 1% bovine serum albumin (BSA, Sigma, A9647), 0.1% Triton (Sigma, T8787)) or, for organoid sections, solution 2 (1X PBS, 2% BSA, 0.1% Tween20 (Sigma, P1379)). RPE1 cells were blocked with 3% BSA in PBS. Coverslips and sections were incubated overnight at 4°C with primary antibodies (Supplementary Table 4) diluted in saturation solution, then washed three times with PBS and incubated with secondary antibodies in the saturation solution for 1h30 at room temperature in the dark. After they were washed thrice with PBS, they were incubated with 100 ng/mL DAPI (Invitrogen, D1306) for 10 minutes and washed. Coverslips were mounted on slides using FluorSave Mounting media (Sigma, 345789), and cryosections Permafluor Aquaous Mounting Media (Fisher Sci., 12695925). Slides were imaged using the Zeiss LSM 800 confocal microscope (Carl Zeiss). All images were analyzed using the Fiji (ImageJ) software. For neural rosette measurements, the areas were quantified by detouring the neural rosette or the lumen. The thickness was assessed by the following formula: (mean of four rosette diameters – mean of four lumen diameters) divided by 2.

### Statistical analysis

All the data are reported as the mean with standard deviation (SD) or median with 95% confidence interval (CI) of at least three independent experiments. Normality of datasets was evaluated using the Shapiro-Wilk test, and outliers identified with the Grubb’s test. All hypothesis tests were two-sided, and statistically significant differences (p-value<0.05) were calculated by parametric or non-parametric tests as indicated in figure legends. Statistical analyses were performed using GraphPad Prism software.

## Results

### Identification of *RTTN* mutation in a patient with TALS-like traits

A sample from a child suspected to have Taybi-Linder syndrome was addressed to us for genetic diagnosis. The patient is the daughter of healthy first-cousin parents from North Africa without any notable medical history reported in the family. The pregnancy was marked by a severe intrauterine growth retardation with brain abnormalities. The baby girl was born at full term with a weight of -4 DS, a height of -4 DS and a head circumference of - 9 DS. Radiographic findings revealed a global ossification delay, with a prominent occiput, 11 pairs of ribs, but no skeletal dysplasia. In her first months of life, she developed severe, diffuse eczema with alopecia. She showed a moderate global developmental delay, and her stature weight growth retardation worsened, while maintaining harmonious morphology and proportions: during infancy, she weighed -4 SD, measured -4.5 SD and had a head circumference (HC) of -10 SD. During chilhood, HC was -10.5 SD. During adolescence, she weighed -1.5 SD and her height -4 SD. She has a low frontotemporal hair implantation, an extremely sloping forehead, a discrete convergent strabismus, and retrognathism. Brain MRI performed during infancy showed microencephaly with short but complete corpus callosum (arrowhead, Fig. 1A), delayed myelination and supratentorial pachygyria (red arrows, Fig. 1B). The cerebellum and brainstem were normal (Fig. 1A). She is currently a young adult and has a neurological follow-up for cognitive impairment and epilepsy treated with valproate, clobazam and olanzapine.

**Figure 1.**
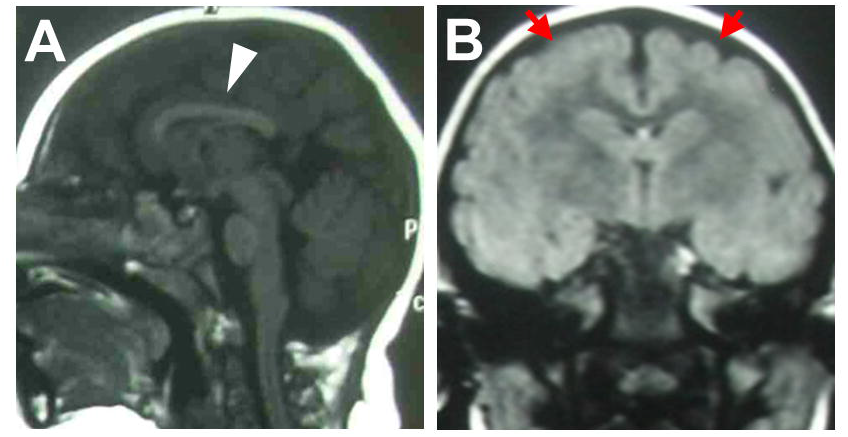
Radiological findings of the TALS-like patient. **(A-B)** Brain MR images on midsagittal T1WI (E) and coronal T2-FLAIR (F) sequences, performed during infancy, showing a short but complete corpus callosum (arrowhead on A) and a supra-tentorial diffuse pachygyria (red arrows on B). Delayed myelination for age was also noted on axial sequences.

Because of the clinical suspicion of TALS, Sanger sequencing of *RNU4ATAC* was first performed, but no variant was found. Then, a whole exome sequencing was done and the homozygous variant c.2953A>G was identified in *RTTN,* a gene that does not contain any minor intron. Interestingly, this case is not the first one of its kind. When we reviewed the literature on cases harboring bi-allelic variants in *RTTN*, two families reported by Grandone *et al.*^21^ and by Shamseldin *et al.*^20^ particularly caught our attention. In the former report, the siblings, a boy and a girl, also homozygous for the c.2953A>G *RTTN* variant, although not described as such, presented typical TALS features including severe growth retardation, severe microcephaly (with lissencephaly), eczema and developmental delay. In the latter, two siblings, negative for *RNU4ATAC* but carrying compound heterozygous missense mutations in *RTTN,* were clinically diagnosed with MOPD1 as they were presenting severe pre- and post-natal growth delay, severe microcephaly (with cerebellar hypoplasia, dysgenesis/agenesis of corpus callosum, lissencephaly or reduced sulcation, ventricular abnormalities), joint contractures and the typical facial dysmorphism (prominent eyes, sloping forehead, micrognathia). The two baby boys died before 3 months of age. Of note, all the other *RTTN* published cases present quite heterogeneous clinical presentations, distinct from the TALS phenotype.

Since the pathophysiological mechanisms by which damaging *RTTN* variants impact brain development are poorly known, we undertook to investigate c.2953A>G further through the study of various cellular models, which will also give us insights for the Taybi-Linder syndrome.

### *RTTN* c.2953A>G variant leads to in-frame exon 23 skipping that is responsible for cell division defects

As previously described,^21,25,30^ the c.2953A>G variant, located at the penultimate nucleotide of exon 23, alters the splicing donor site sequence of intron 23. We first confirmed by RT-PCR, RT-qPCR and RNA sequencing that c.2953A>G induces partial skipping of exon 23 in patient fibroblasts (Supplementary Fig. 1A-D). Indeed, the variant results in the expression of three different RNA isoforms: the full length (FL) isoform, which encodes the amino-acid change p.Arg985Gly (RG); the most prevalent isoform, resulting from the skipping of the in-frame exon 23, which encodes a Rotatin protein depleted of 23 amino-acids (Δ23); and the third isoform, resulting from the skipping of both exons 22 and 23 (Δ22-23), which contains premature stop codons. This latter isoform is partially degraded by nonsense-mediated mRNA decay (Supplementary Fig. 1D), and is likely to give rise to small amounts of a nonfunctional truncated protein that lacks two third of its full length. Globally, the alteration of *RTTN* pre-mRNA splicing does not alter its total level of expression (Supplementary Fig. 1E). To investigate the consequences of *RTTN* c.2953A>G at the protein level, we performed ultrastructural expansion microscopy (U-ExM), allowing precise nanoscale mapping of proteins. We first confirmed in control cells that Rotatin is located at the proximal end of centrioles, lining the internal microtubule wall in both mature and pro-centrioles, and has a ring-like localization from top-viewed centrioles (Supplementary Fig. 1F, Fig. 2A). Rotatin is also present at the basal body, without any signal detected in the axoneme of primary cilia (Supplementary Fig. 1F). In patient cells, we observed a reduced quantity of Rotatin at the centrioles (Fig. 2A, B), suggesting a defective targeting or recruitment of mutated Rotatin, which was accompanied by a slight decrease of centriole length (Fig. 2C). Of note, and contrary to what was previously published,^30^ we observed no alteration of cell cycle progression and mitosis events in patient fibroblasts (Supplementary Fig. 2A-D).

**Figure 2.**
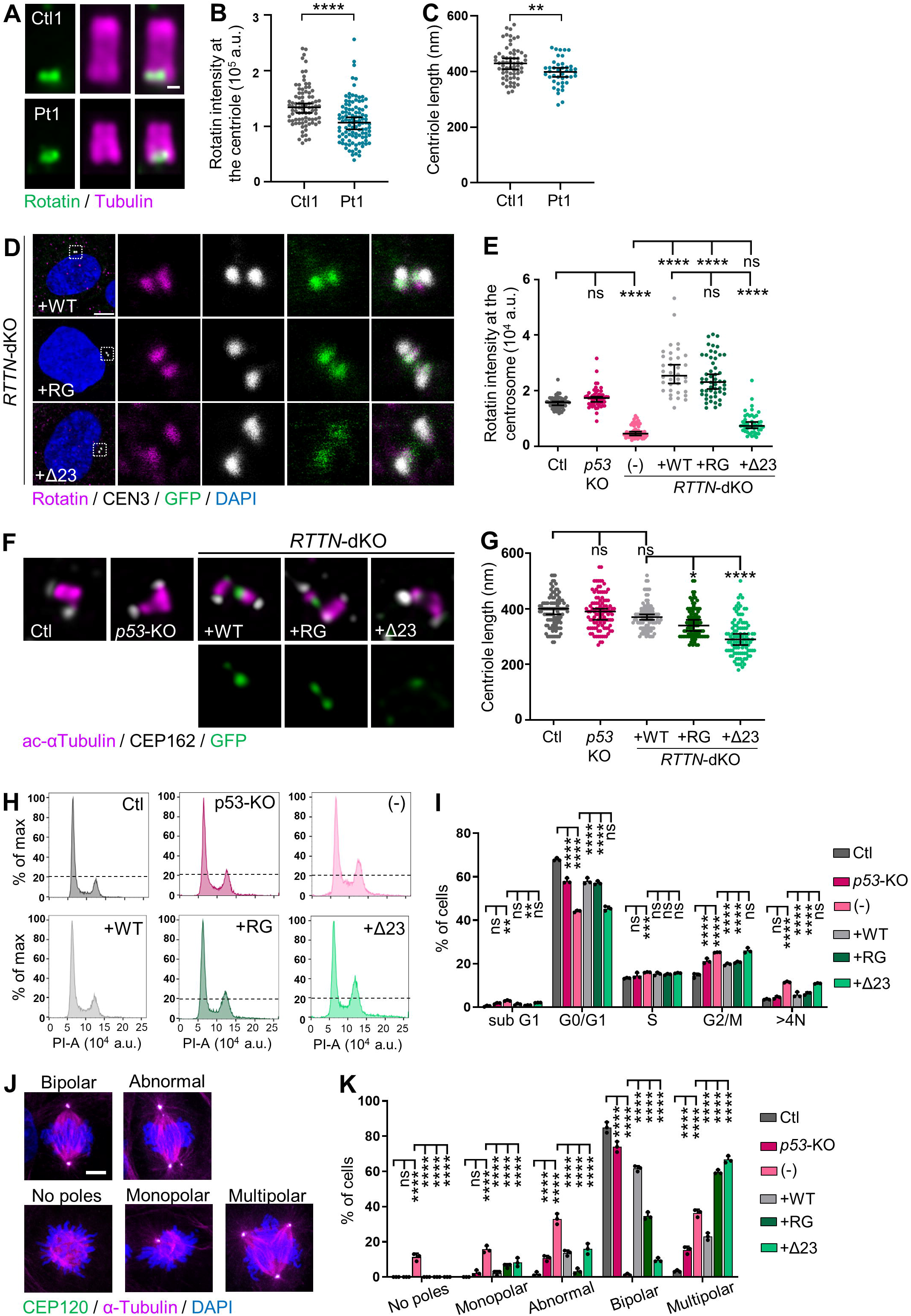
*RTTN* c.2953A>G variant alters Rotatin localization at centriole, cell cycle progression and mitosis in *RTTN*-dKO RPE1 cellular model. Experiments were performed in control (Ctl1) and patient (Pt1) fibroblasts (A-C), or in control, *p53*-KO and *RTTN*-dKO RPE1 cells induced to express wild-type (WT) or mutated (RG, Δ23) RTTN-GFP proteins (D-K). **(A)** Representative confocal images of Rotatin (green) in expanded centrioles, labeled with tubulin (magenta), in fibroblasts. **(B, C)** Quantification of Rotatin intensity at the centrioles (B) and of length of centrioles (C), such as shown in A. Graphs show the median ± 95% CI from three independent experiments (n>85 (B) or n>40 (C) centrioles). **(D)** Representative confocal images of Rotatin (magenta) localisation at the centrosome in *RTTN*-dKO RPE1 cellular model. Centrin3 (CEN3, grey) labels centrosomes. **(E)** Quantification of Rotatin intensity, relative to CEN3, at the centrioles, such as shown in D and in Supplementary Fig. 2A. Graph shows the median ± 95% CI from three independent experiments (n>35 centrosomes). **(F)** Representative super-resolution images of centrioles in *RTTN*-dKO RPE1 cellular model. Acetylated α-Tubulin (magenta) labels centriole structure and CEP162 its distal end. **(G)** Quantification of length of centrioles, such as seen in F. Graph shows the median ± 95% CI from three independent experiments (n≥100 centrioles). **(H)** Flow cytometric cell cycle analysis histograms of *RTTN*-dKO RPE1 cellular model. The dotted line represents the top of G2/M peak in Ctl cells for reference. **(I)** Quantification of the percentage of cells in each cell cycle phase, such as seen in H. Graph shows the mean ± SD of three independent experiments. **(J)** Representative confocal images of metaphases seen in *RTTN*-dKO RPE1 cellular model. Acetylated α-Tubulin labels mitotic spindles and CEP120 centrosomes. **(K)** Quantification of the proportion of cells with the indicated mitotic phenotypes. Graph shows the mean ± SD of three independent experiments (n ≥ 100 cells per experiment). ns, not significant; *p-value<0.05; **p-value<0.01; ***p-value<0.001; ****p-value<0.0001 following Mann-Whitney test (B), t-test (C), Kruskal-Wallis with Dunn’s Multiple Comparison test (E, G) or two-way ANOVA with Tukey’s correction (I, K). Scale bars: 100 nm (A), 5 µm (D). DAPI stains DNA. a.u. arbitrary units, PI propidium iodide.

To evaluate the respective effect of the missense p.Arg985Gly and Δ23 mutations, we infected the previously described double knock-out *p53^-/-^;RTTN^-/-^* (*RTTN*-dKO) RPE1 cells^35^ with each of the two doxycycline-inducible GFP-tagged RTTN isoforms. We first verified that they were correctly expressed by immunofluorescence and western blot (Supplementary Fig. 3A, B, Fig. 2D). Contrary to WT and RG Rotatin proteins which both localize to mature and pro-centrioles, we observed that the overall Δ23 Rotatin abundance at centrioles is low (Fig. 2D, E). As a consequence, loading of downstream centriolar proteins such as POC1B^34^ is altered in presence of Δ23 Rotatin (Supplementary Fig. 3C, D). Following the decreased localization of both Rotatin and POC1B to centrioles, we observed a shortening of centrioles in *RTTN*-dKO RPE1 cells expressing Δ23 Rotatin (Fig. 2F, G). We also observed a slight G2/M-phase retention and aneuploidy (Fig. 2H, I), as well as an increase of abnormal mitotic events (Fig. 2J, K), in *RTTN*-dKO cells induced to express Δ23 Rotatin compared to the WT form (Fig. 2H-K).^35^

Hence, taken together, these results show that the protein Δ23 Rotatin has the most deleterious effect, and that the loss of 23 amino-acids has not only a deleterious impact on the targeting or recruitment of Rotatin to centrioles, but that it also alters its function in recruiting downstream effectors of centriole maturation. These alterations may explain the defective division of cells.

### *RTTN* c.2953A>G variant disrupts cilium formation and function

As aforementioned, Rotatin localizes to the base of primary cilia in quiescent cells (Supplementary Fig. 1F) and previous reports showed that primary cilium length was significantly reduced in some patient fibroblasts.^19,24,30^ Hence, we investigated the formation, disassembly and function of primary cilia as a consequence of c.2953A>G.

First, and contrary to what has been previously reported for other *RTTN* mutations,^19,24,30^ we did not observe any alterations of proportion of ciliated cells or cilium length in the patient fibroblasts (Fig. 3A-C). However, in *RTTN*-dKO RPE1 cells, we did observe a drastic loss of primary cilium formation, that was partially rescued by the WT and RG Rotatin forms (both count and length) but not by the Δ23 variant (Supplementary Fig. 4A-C). We thus concluded that Rotatin is essential for cilium formation – as it was shown by RNA interference^19^ – but that the *RTTN* c.2953G>A variant in patient fibroblasts may produce enough RG protein to allow cilium formation.

**Figure 3.**
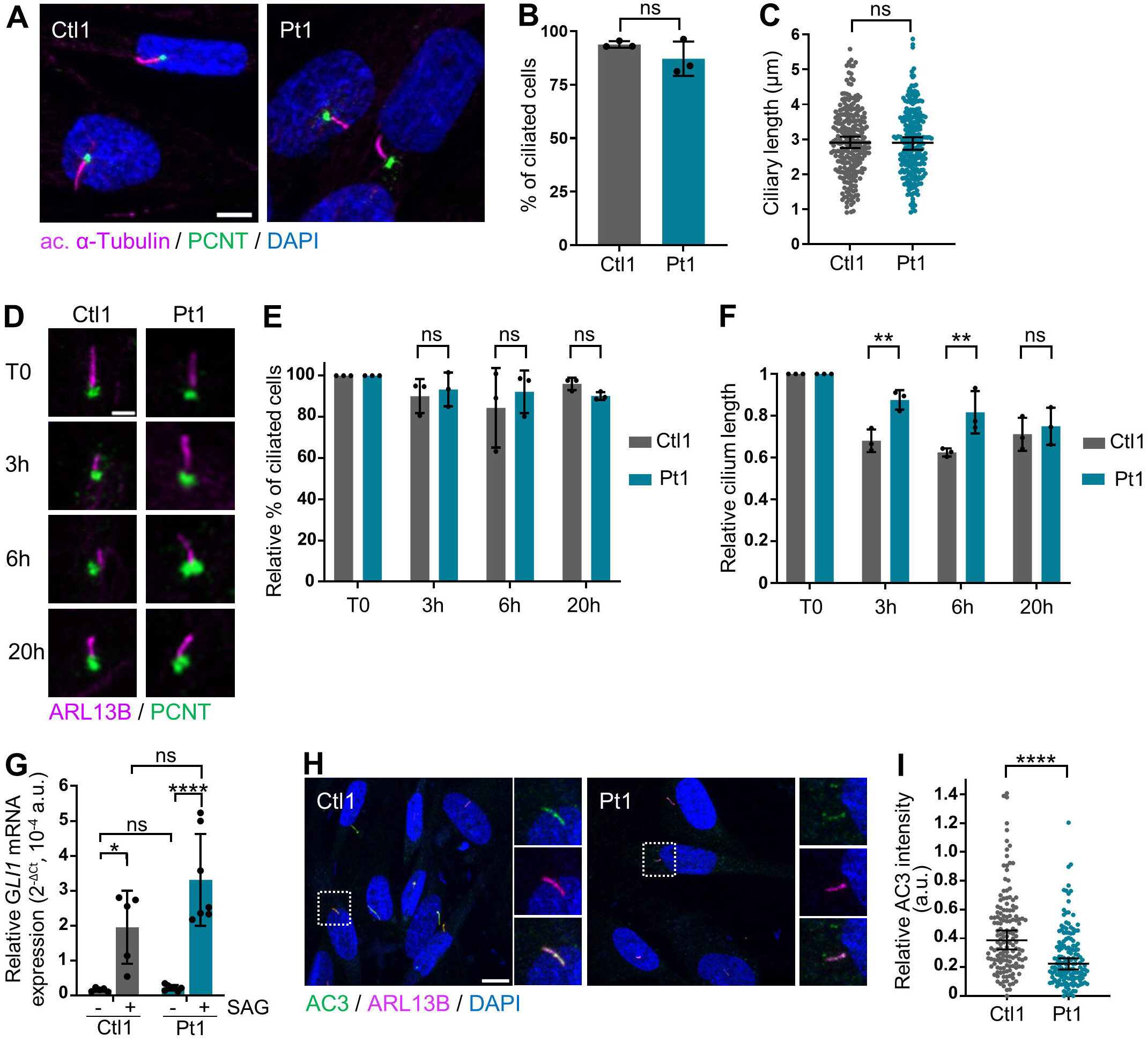
*RTTN* c.2953A>G variant alters ciliary function in patient fibroblasts. All experiments were performed in control (Ctl1) and patient (Pt1) fibroblasts. **(A)** Representative confocal images of primary cilium (acetylated α-Tubulin, magenta) and centrosome (PCNT, green) in fibroblasts. **(B, C)** Quantification of percentage of ciliated cells (B) and of length of cilia (C) observed in A. Graphs show the mean ± SD (B) or median ± 95% CI (C) of three independent experiments (n=250 cells). **(D)** Representative confocal images of disassembly of primary cilium (ARL13B, magenta) in fibroblasts, at serum starvation (T0) or at T=3, 6 or 20 hours after serum addition in the culture medium. PCNT (green) labels centrosome. **(E, F)** Quantification relative to T0 of percentage of ciliated cells (E) and of length of cilia (F) observed in D. Graphs show the mean ± SD of three independent experiments (n>100 cells). **(G)** RT-qPCR analysis of *GLI1* relative expression in fibroblasts not treated or treated with SAG. *ACTB* was used as house-keeping gene. Graph shows the mean ± SD of five independent experiments. **(H)** Representative confocal images of adenylate cyclase 3 (AC3, green) in primary cilium (ARL13B, magenta) in fibroblasts. **(I)** Quantification of ciliary AC3 staining intensity relative to ARL13B. Graph shows the median ± 95% CI of three independent experiments (n=150 cells). ns, not significant; *p-value<0.05; **p-value<0.01; ***p-value<0.001; ****p-value<0.0001 following Mann-Whitney test (B, C, I), two-way ANOVA with Bonferroni’s correction (E, F), two-way ANOVA with Tukey’s correction for multiple tests (G). Scale bars: 5 µm (A), 2 µm (D), 10 µm (H). DAPI labels nuclei. a.u. arbitrary units.

Next, we performed a dynamic analysis of cilium disassembly in patient fibroblasts. After promoting cilium formation for 48 hours by serum starvation, ciliary disassembly was induced upon serum addition. While the percentage of ciliary cells was unchanged in both control and patient cells, we observed a decrease of cilium length in control cells from T=3 hours following serum addition that was less pronounced in patient cells (Fig. 3D-F), suggesting that loss of function of Rotatin may lead to a delayed cell cycle re-entry, which is an important parameter in highly proliferative cells, such as neural stem cells.^37^

Finally, we investigated primary cilium function by analyzing two ciliary signaling pathways. The first one is the Sonic Hedgehog (Shh) pathway, in which the binding of the ligand Shh to its ciliary receptor Ptch1 alleviates the inhibitory signal that maintains Smoothened (Smo) out of the cilium. Upon Shh activation, Smo translocates into the cilium and activates the processing machinery of Gli transcription factors. Once cleaved, these factors shuttle into the nucleus to control Shh target gene expression (including *GLI1*). Upon treatment with the Smo agonist SAG to activate the Shh pathway, we observed an upregulation of *GLI1* expression in control cells, which is slightly higher in patient fibroblasts (albeit not significantly) (Fig. 3G). As a second ciliary signaling pathway, we investigated the ciliary localization of the adenylate cyclase III (AC3) enzyme that locally generates cAMP from ATP and participates, among others, to the control of cilium length and Shh pathway.^39^ We observed a significant decrease in AC3 ciliary localization in patient fibroblasts (Fig. 3H-I), thus suggesting that Rotatin is required for the proper ciliary localisation of AC3 and other signaling molecules to ensure cilium function.

Altogether, these results show that Rotatin is required for both cilium formation and function, and that Δ23 Rotatin is once again the most deficient form.

### *RTTN* c.2953A>G mutation leads to cell cycle defects, abnormal mitotic events and increased cell death in iPSC-derived neuronal stem cells

To be able to better investigate the pathophysiological mechanisms involved in the microcephaly and brain abnormalities seen in the patient, we generated CRISPR/Cas9-edited induced pluripotent stem cells (iPSC) (Supplementary Methods, Supplementary Fig. 5A-D). In iPSC, the c.2953A>G variant had similar impact on *RTTN* pre-mRNA splicing as that seen in fibroblasts, except that the Δ23 isoform was more abundant (Supplementary Fig. 5E, F); as in fibroblasts, the levels of mRNA expression were unchanged (Supplementary Fig. 5G). A comprehensive analysis of CRISPR/Cas9-edited iPSC phenotypes showed no significant differences between WT and *RTTN* mutated (KI) clones regarding cell cycle progression (Supplementary Fig. 6A, B), mitotic events (Supplementary Fig. 6C, D), percentage of ciliated cells and cilium length (Supplementary Fig. 6E-G).

The iPSC were then differentiated into neuronal stem cells (NSC) (Supplementary Fig. 7A-C). In this cell type, the proportion of the three mRNA isoforms resulting from the presence of c.2953A>G was as seen in fibroblasts (Supplementary Fig. 7D-F). No strong alteration of centriole length was detected in KI NSC (Supplementary Fig. 7G, H), neither significant abnormalities regarding the percentage of ciliated cells, cilium length or function (as far as AC3 ciliary localization is concerned) (Supplementary Fig. 8A-E); however, we observed by flow cytometry a higher proportion of KI NSC with aneuploidy and with retention in G2/M phase at the expense of phase G1 compared to WT NSC (Fig. 4A, B). Careful analysis of mitosis events by immunofluorescence allowed the detection of various defects of the mitotic spindles in KI NSC: whereas in WT NSC, 90% of mitosis were normal with 2 spindle poles at the metaphase, more than 40% of mitosis in KI NSC showed anomalies, such as no poles, one pole, asymmetric poles or multiple poles (Fig. 4C, D), thus confirming the observation seen by flow cytometry. In order to determine the fate of the cells with defective mitosis, we investigated markers of cell arrest and apoptosis. We observed a higher number of double-positive p53+; p21+ cells in KI NSC compared to controls, as well as an increase of cleaved-Caspase 3 staining (Fig. 4E-H), indicating that a higher proportion of KI NSC undergo cell cycle arrest and/or apoptosis. Then, to further explore mitotic events, we investigated spindle orientation by determining the angle formed by the two spindle poles.^35^ Whereas in most WT NSC, the two centrosomes were aligned within a plane parallel to the support (between 5 and 10°), they were slightly misaligned in most KI NSC (15-30°) (Fig. 4I, J). This result suggests that a higher proportion of KI NSC could be prone to undergo asymmetric divisions and thus to prematurely shift to neurogenic cell divisions, which, as a result, would exhaust the progenitor pool.^36,40,41^

**Figure 4.**
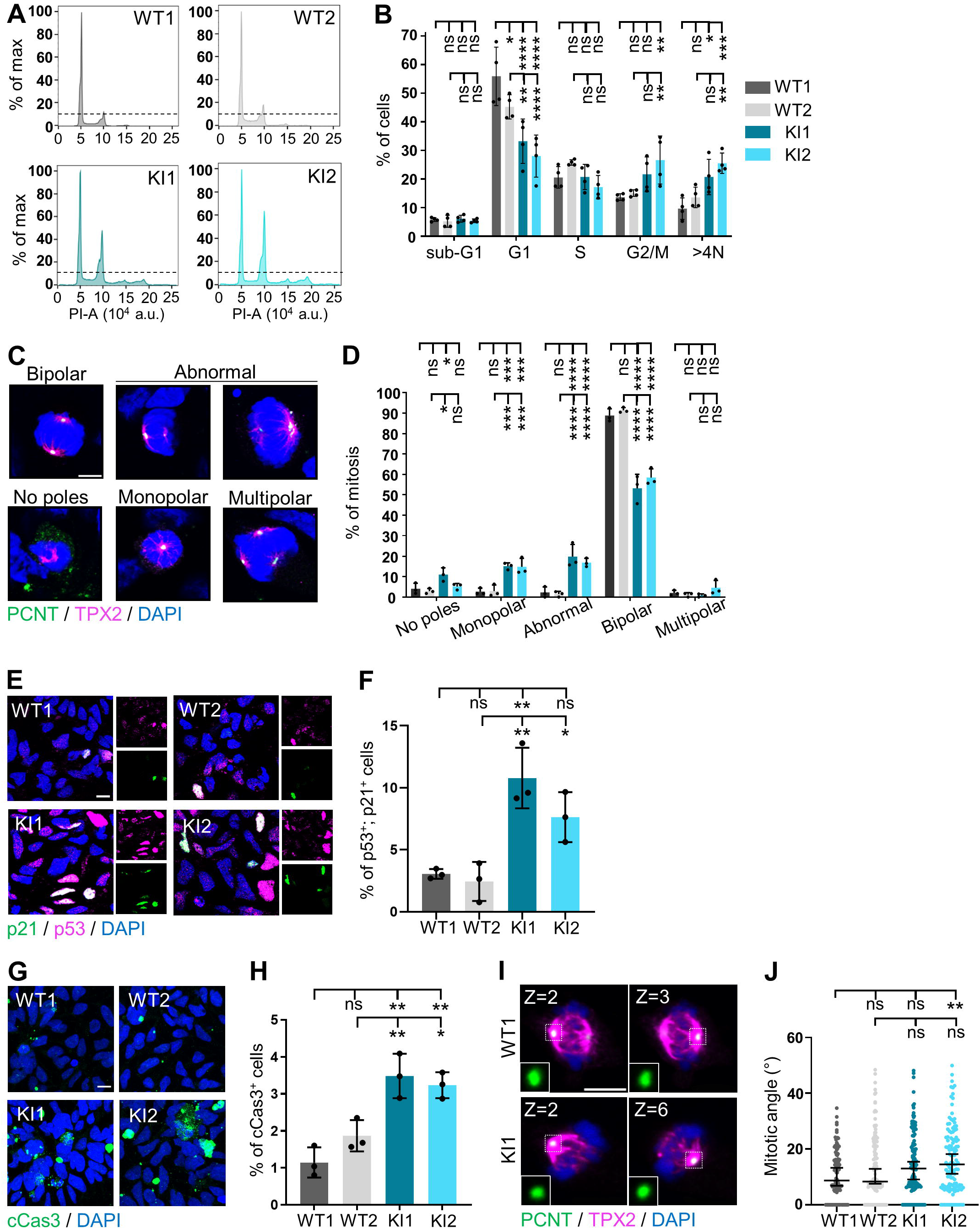
*RTTN* c.2953A>G variant alters mitosis and cell cycle progression in neural stem cells. All experiments were performed in control (WT) and *RTTN*-mutated (KI) NSC. **(A)** Flow cytometric cell cycle analysis histograms in NSC. The dotted line marks the maximal value of G2/M peak in WT1 cells for reference. **(B)** Quantification of the percentage of cells in each cell cycle phase. Graph shows the mean ± SD of three independent experiments. **(C)** Representative confocal images of metaphases observed in NSC. PCNT (green) labels the centrosomes, TPX2 (magenta) the mitotic spindle. **(D)** Quantification of the proportion of cells with the indicated mitotic phenotypes. Graph shows the mean ± SD of three independent experiments (n>100 mitosis per experiment). **(E)** Representative confocal images of cells stained with p53 (magenta) and p21 (green), markers of cell arrest in NSC. **(F)** Quantification of the percentage of p53; p21-double positive cells, such as seen in E. Graph shows the mean ± SD of three independent experiments (n>800 cells per experiment). **(G)** Representative confocal images of apoptotic cells (cleaved Caspase-3, green) in NSC. **(H)** Quantification of the percentage of cCas3-positive cells, such as seen in G. Graph shows the mean ± SD of three independent experiments (n>800 cells per experiment). **(I)** Representative confocal images of metaphases in NSC. PCNT (green) labels the centrosomes, TPX2 (magenta) the mitotic spindle. In insets are shown each of the two the spindle poles located in the indicated Z-plane. **(J)** Quantification of the mitotic angle based on the distance and the height between both spindle poles. Graph shows the median ± 95% CI from three independent experiments. ns, not significant; *p-value<0.05; **p-value<0.01; ***p-value<0.001; ****p-value<0.0001 following two-way (B, D) or one-way (F, H) ANOVA with Tukey’s correction, or Kruskal-Wallis with Dunn’s correction (J). Scale bars: 5 µm (C, I), 10 µm (E, G). DAPI labels nuclei. a.u., arbitrary units.

### *RTTN* c.2953A>G*-*mutated cortical organoids display growth defects with delayed neural rosette formation and increased cell death

We generated matrix-free cortical organoids (CO) derived from CRISPR/Cas9-edited iPSC (Supplementary Fig. 9A-C), in order to mimic in three dimensions the early stages of brain development. Indeed, the organization of NSC into neural rosettes resembles the developmental structures of neural tubes. First, in order to check if the splicing outcomes of *RTTN* pre-mRNA carrying c.2953A>G was modified in the context of an increased cellular complexity, we analyzed the RNA isoforms in days in vitro 35 (DIV35) CO. We found the same three isoforms as before, in the same proportions as in fibroblasts and NSC, with variations in total *RTTN* mRNA expression not correlated with the genotypes (Supplementary Fig. 9D-F).

We then monitored CO growth by measuring their size twice a week, over a period of culture of 67 days. We observed that starting at DIV21, KI CO exhibited a slower growth rate compared to WT CO, being more than half smaller at DIV67 (Fig. 5A, B). In addition to alteration of CO size, we noticed that KI CO morphology was altered: while neural rosettes were appearing at the periphery in WT CO as early as DIV35, these structures were seen in KI CO from DIV56 onwards only (Fig. 5A, arrowheads).

**Figure 5.**
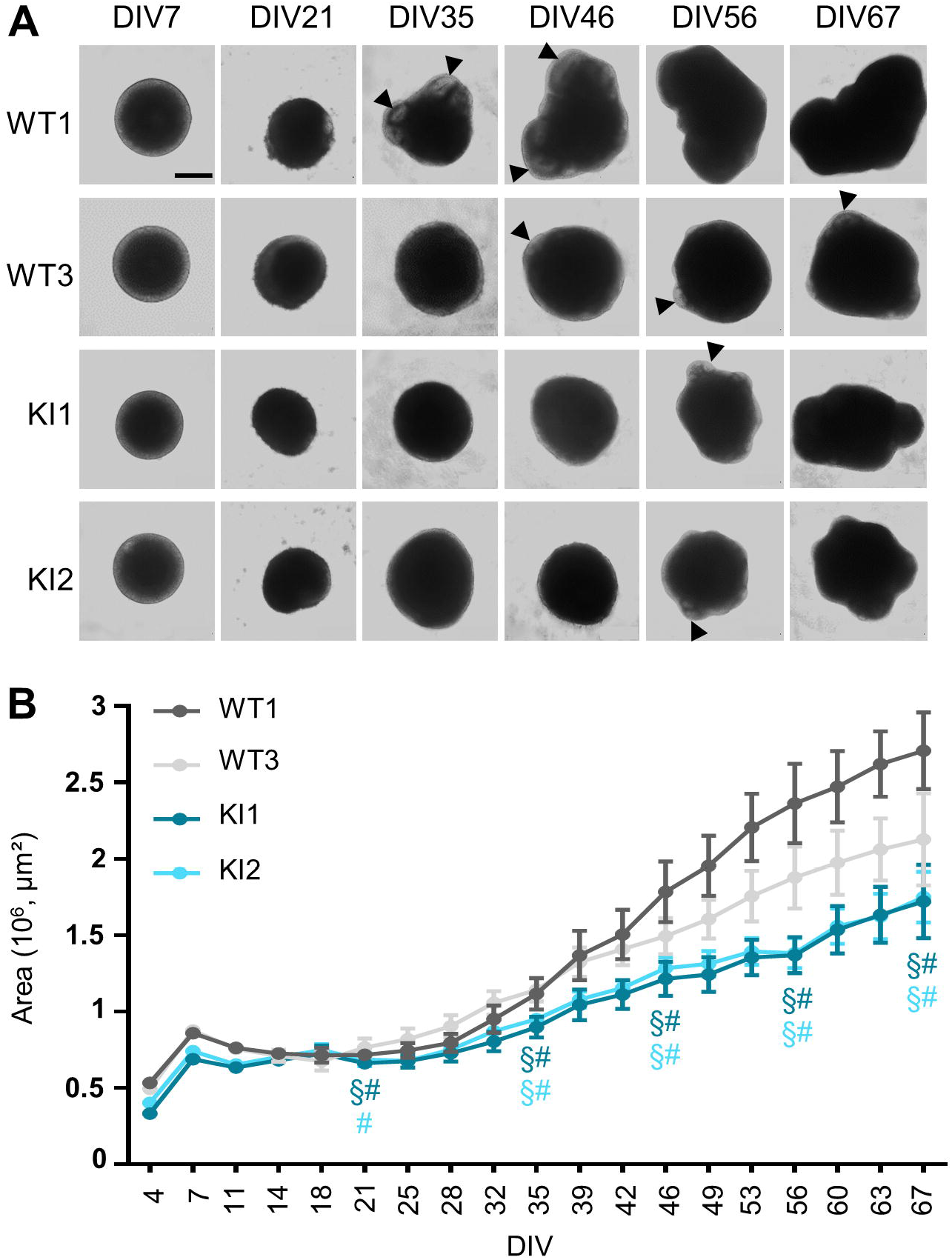
*RTTN* c.2953A>G variant alters cortical organoid growth. All experiments were performed in wild-type (WT) and *RTTN*-mutated (KI) cortical organoids (CO). **(A)** Representative optical images of CO at different time points, from DIV7 to DIV67. Arrowheads indicate neural rosettes seen in transparency. **(B)** Growth curve of CO such as seen in A. Graph shows the mean ± SD of 20 CO areas at each time point. Statistical analyses are shown for DIV21, DIV35, DIV46, DIV56 and DIV67 only. § and # indicate a p-value<0.001 when compared to WT1 and WT3 respectively, following two-way ANOVA with Tukey’s multiple correction. Scale bar: 500 nm. DIV, days in vitro.

Next, to have a better understanding of neural rosette formation in the context of the presence of c.2953A>G, we performed immunostaining on CO cryosections, using SOX2 to label NSC and N-cadherin to label their apical adherent junctions facing the rosette lumen. By quantifying both their number and size, we found that they were fewer and smaller in KI CO from DIV35 to DIV56 (Fig. 6A-C). KI neural rosettes seemed also less well-organized, with SOX2+ cells less elongated and less stratified in the rosette width, as indicated by the decreased thickness of neural rosettes, while rosette lumen area remained unchanged (Supplementary Fig. 9G, H). We thus concluded from these experiments that the presence of c.2953A>G in *RTTN* leads to the delayed formation of neural rosettes. In accordance to that, we noticed that the number of N-cadherin-positive foci, a mark of NSC polarity within neural rosettes, was lower in KI CO at DIV35 compared to controls (Fig. 6A, D). This finding was further confirmed by the study of two other NSC apical markers, i.e. ZO-1 (tight junctions) and ARL13B (cilia) (Fig. 6E-F). Hence, these results support the hypothesis that delayed formation of neural rosettes in KI CO is probably due to alteration of NSC polarization.

**Figure 6:**
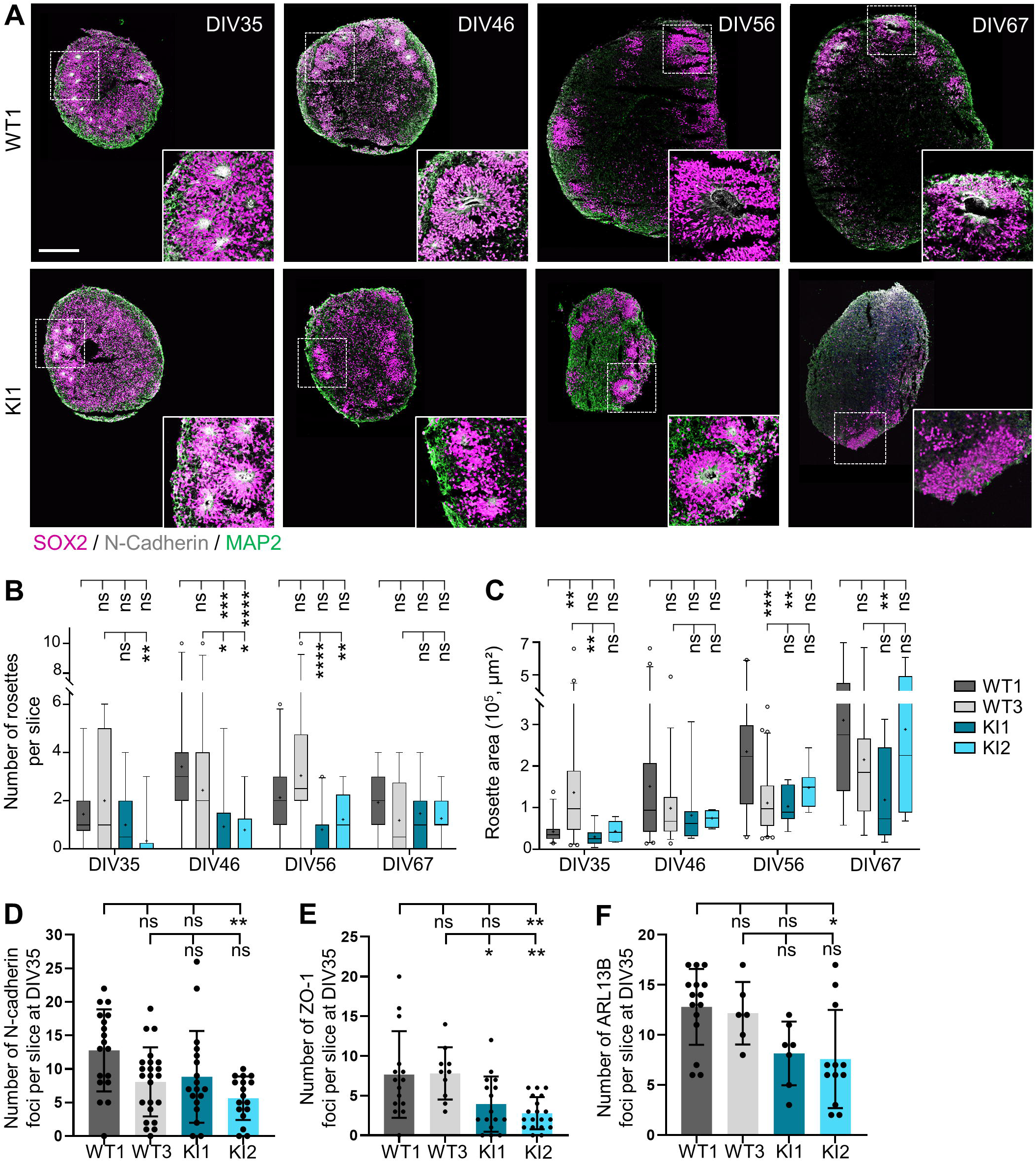
*RTTN* c.2953A>G variant impedes the formation of neural rosettes in the cortical organoids. All experiments were performed in wild-type (WT) and *RTTN*-mutated (KI) cortical organoids (CO). **(A)** Representative confocal images of neural rosettes in CO sections at DIV35, DIV46, DIV56 and DIV67. SOX2 (magenta) labels neural stem progenitors (NSC), N-cadherin (grey) the rosette lumen, and MAP2 (green) neurons. **(B-C)** Quantification observed overtime of the number (B) and area (C) of the rosettes, such as seen in A. Box-and-whisker plots display, in the box, the median (middle bar), the mean (cross) and the 25th-75th percentiles, and in whiskers, the 5th to 95th percentile of values from 4 different organoids (n=6 to 20 slices in total). **(D-F)** Quantification of the number of N-cadherin (D), ZO1 (E) and ciliary (F) foci observed in CO at DIV35. N-Cadherin is a marker of the adherent junctions (as seen in A), ZO1 the tight junctions, and ARL13B the cilium. Graphs show the mean ± SD from 4 different CO (n=6 to 20 slices total). ns, non-significant; *p-value<0.05; **p-value<0.01; ***p-value<0.001; ***p-value<0.0001 following two-way ANOVA with Tukey’s correction (B, C) or Kruskal-Wallis with Dunn’s multiple comparisons test (D-F). Scale bar: 250 µm. DIV, days in vitro.

Finally, to investigate whether a defect in cell proliferation could also be involved in the decreased rosette number and size observed in *RTTN*-mutated CO, we performed immunostainings with Ki67 (Fig. 7A). Globally, at DIV35, we observed that the decreased number of SOX2+ cells in KI CO compared to controls (Fig. 7A, B) was concomitant with a decrease of Ki67 staining (Fig. 7A, C), thus suggesting an overall lower number of cycling NSC in *RTTN*-mutated CO. We further analyzed NSC divisions that occur at the center of neural rosettes, and performed immunostainings to label dividing NSC with TPX2, a marker of spindles, and phospho-Vimentin (Fig. 7E). We observed that the number of mitotic events per rosette was decreased in KI CO at DIV35 and DIV46 (Fig. 7E, F). Finally, by analyzing cell apoptosis, we observed an increase of cleaved caspase-3 staining in KI neural rosettes compared to controls, at both DIV35 and DIV56 (Fig. 7D). It is noteworthy that alterations of SOX2+ cell number, Ki67 positivity or number of mitotic events tend to decline at late time points (DIV56), to be even reversed at DIV67 for mitotic events (Fig. 7B, C, F).

**Figure 7:**
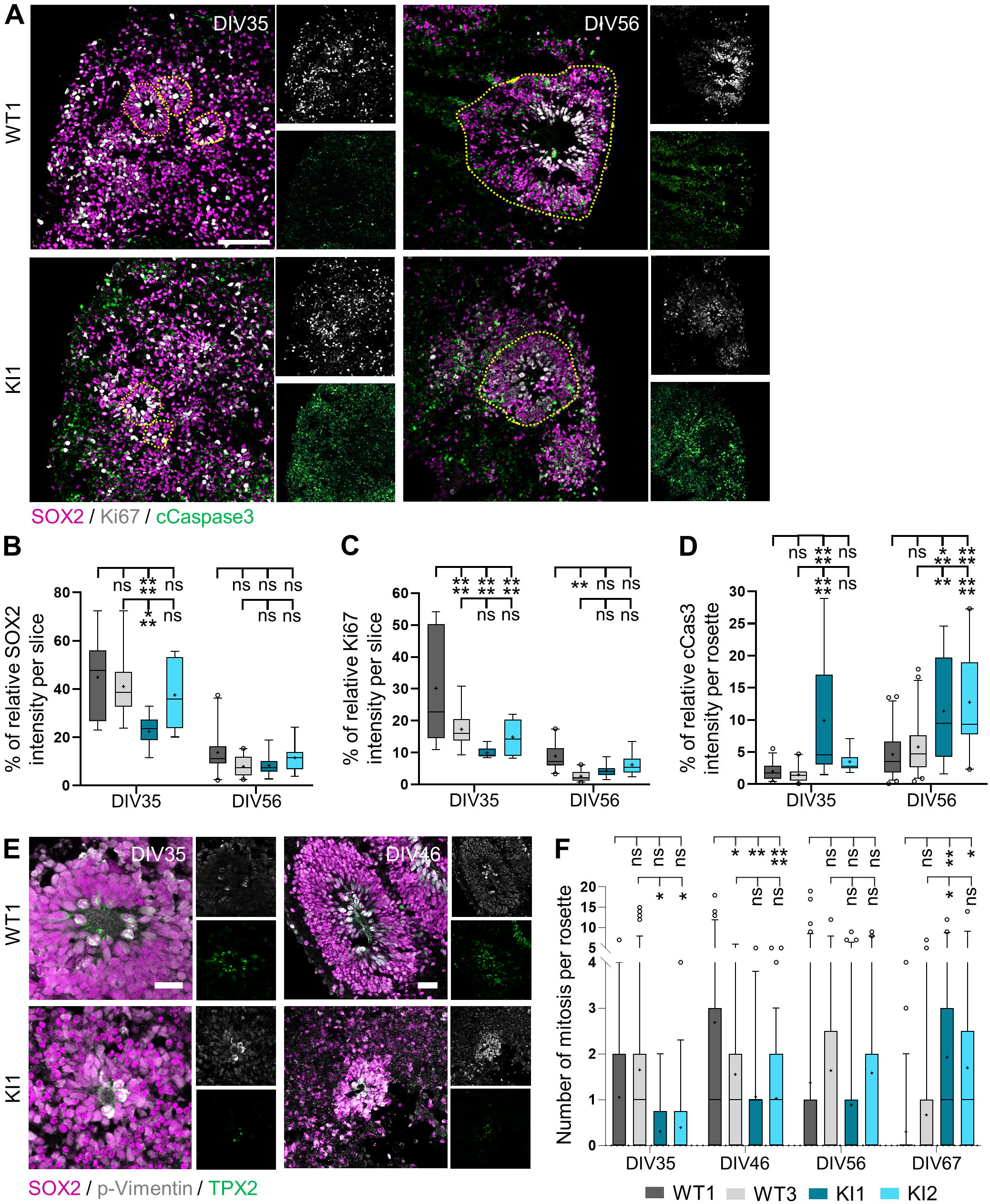
*RTTN*-mutated organoids exhibit decreased proliferation and increased cell death at early stages. All experiments were performed in wild-type (WT) and *RTTN*-mutated (KI) cortical organoids (CO). **(A)** Representative confocal images of proliferative (Ki67, grey) and apoptotic (cleaved Caspase-3, green) cells in CO at DIV35 and DIV56. SOX2 (magenta) labels neuronal progenitor cells. Neural rosettes are highlighted by yellow dashed lines. **(B-D)** Quantification of the percentage of SOX2+ (B), Ki67+ (C) and cCaspase-3+ (D) cells such as seen in A, based on the relative intensity of the markers towards DAPI. Box-and-whisker plots display, in the box, the median (middle bar), the mean (cross) and the 25th-75th percentiles, and in whiskers, the 5th to 95th percentile of values from 4 different CO (n=6 to 20 slices in total). **(E)** Representative confocal images of dividing cells (pVimentin, grey; TPX2, green) in WT and KI neural rosettes at DIV35 and DIV56. SOX2 (magenta) labels neural progenitors. **(F)** Quantification of mitotic events observed in neural rosettes such as seen in E. Box-and-whisker plots display, in the box, the median (middle bar), the mean (cross) and the 25th-75th percentiles, and in whiskers, the 5th to 95th percentile of values from 4 different CO (n=6 to 20 slices total). ns, non-significant; *p-value<0.05; **p-value<0.01; ***p-value<0.001; ****p-value<0.0001 following two-way ANOVA with Tukey’s correction (B-D, F). Scale bars: 100 µm (A), 50 µm (E).

Hence, altogether, these data suggest that the alteration of several cellular processes, including NSC polarization, division and survival, contribute to the delay of rosette maturation in *RTTN*-mutated CO.

## Discussion

In this article, we investigated the pathophysiological mechanisms underlying the pathogenicity of the *RTTN* c.2953A>G variant identified in a patient with a Taybi-Linder-like syndrome, in fibroblasts derived from the patient, and for the first time, in human iPSC-derived neuronal models. We showed that this variant mostly exerts its pathogenicity by inducing the skipping of in-frame exon 23 that encodes 23 amino-acids, and that it has a pleiotropic impact on diverse cellular processes occurring concomitantly or in cascade, including alterations of cell organization/polarity, division and survival of neuronal progenitors. All these alterations contribute to NSC pool reduction, thus leading to the microcephaly seen in the patient we describe here and in most *RTTN* mutation carriers.

The *RTTN* c.2953A>G variant is present in a total of 5 out of 38 reported cases (in 4 out of 23 families), who all originate from North Africa, thus suggesting a founder effect.^21,25,30^ It has been reported in the gnomAD database at the heterozygous state in 4 individuals only (out of 725,468 sequenced), all of them not unambiguously clustering with the major populations in a principal component analysis. It is noteworthy that it is the only recurrent *RTTN* variant, all the other reported patients carrying private mutations. Whereas in previous reports it was suspected that patient symptoms resulted from the combined effects of the expression, at the protein level, of both the missense p.Arg985Gly and the p.Ser963_Arg985del (Δ23) Rotatin forms,^21,25,30^ here we show that the missense p.Arg985Gly has barely no pathogenicity by itself. Indeed, it rescues, as efficiently as the WT form, all tested phenotypes associated to the loss of Rotatin in *RTTN*-dKO RPE1 cells. This result is somehow surprising since all algorithms predicting pathogenicity of missense variants classify p.Arg985Gly as pathogenic (scores for Polyphen: 0.977; CADD: 29.6; AlphaMissense: 0.6). This underscores the fact that even if prediction tools are becoming more and more efficient, they remain predictions; functional evaluation is the only way to provide trustworthy insight into variant pathogenicity. In addition to bringing information about the absence of a damaging effect of the p.Arg985Gly variant *per se*, our study highlights here the crucial role of the 23 amino-acids (Ser963 to Arg985) that yet, do not encompass a known functional domain. These amino-acids are located outside the interacting domain with STIL (1-889 aa) that recruits Rotatin to centrioles.^34^ Hence, we hypothesize that the deletion of the 23 amino-acids may alter its conformation and/or the interaction with Rotatin partners – other than STIL – that help it to be brought to the centrosome.

It is interesting to note that the levels of expression of the three isoforms do not vary much in the various cellular models used in this study (skin fibroblasts, iPSC, NSC and cortical organoids), and yet, we do not observe in each of these models all the cellular alterations we describe in total. No alterations were seen in iPSC, which might be explained by their tolerance to mutations and cellular abnormalities that preserves the stem cell physiology integrity.^42^ Only slight alterations of cilium disassembly and ciliary function were observed in patient fibroblasts, while no defects of cell cycle and cell division were noticed. We could argue here that fibroblasts are differentiated cells that do not highly proliferate; nevertheless, our results contradict previous published work,^30^ which highlights the issue of individual/clonal variability. Not surprisingly, given the severe brain malformations associated with *RTTN* mutations, the most striking phenotypes were detected in the neuronal progenitor cells, cultured in 2D or 3D. In this cell type, we demonstrated a high retention of NSC in G2-M phase with numerous mitotic abnormalities that likely results in aneuploidy, cell arrest or cell apoptosis. We also observed a misorientation of the mitotic spindles that may favour a premature differentiation of NSC into neurons. All these alterations of NSC were also seen in cortical organoids (decreased proliferation/cell division, increased apoptosis), and are thus likely to contribute to the reduction of the NSC pool, which is responsible for the reduced number and size of neural rosettes and hence, of organoids, mimicking the microcephaly seen in the patient.

Further, the use of cortical organoid model allowed us to discover a new role of Rotatin in the self-organization of NSC into neural rosettes. Indeed, we observed fewer apical foci (positive for ZO-1, N-cadherin, and primary cilium) in *RTTN*-mutated organoids, suggesting a defect in apico-basal polarity establishment of the bipolar NSC. Neural rosette formation is based on five different steps: cell intercalation, constriction, polarization, elongation and lumen formation.^43^ Once formed, rosettes continue their expansion by fusing with each other.^44^ The molecular mechanisms involved in these steps are not fully understood; they include various factors such as actin cytoskeleton remodeling, calcium, FGF2 or BMP signalings.^43–46^ The question remains how the centrosome/primary cilium complex contributes to these steps. We showed here that Rotatin is rather involved in ciliary function than cilium formation, even though in RPE1 cells the total depletion of *RTTN* leads to absence of primary cilia. Hence, we can hypothesize that misregulation of ciliary signaling, deriving from chemical or mechanical cues, could alter NSC organization into neural rosettes in absence of functional Rotatin. It is noteworthy to remind here that we recently established a link between *U4atac* and primary cilium functions.^47^ Not only does it reinforce the possible feature overlap between *RNU4ATAC* and *RTTN* mutated patients, but it also stresses the importance of primary cilium defects in the pathophysiology of primary microcephaly syndromes – even though this clinical feature is not among those of ciliopathies.

Altogether, through the study of the consequences of the only recurrent *RTTN* variant, we shed light on the pleiotropic functions of Rotatin, whose central role in brain development is starting to emerge.

## Data Availability

All data produced in the present study are available upon reasonable request to the authors

## Acknowledgements

We thank the family and the patient for their contribution to this project, as well as Dr Ester Zuazo Zamalloa (Zumarraga Hospital, Gipuzkoa, Spain) for addressing the patient to us and transferring clinical data. We thank the Centre de Biotechnologie Cellulaire Biotec biobank for biosample management (Emilie Chopin, Isabelle Rouvet), Dr Gaetan Lesca for exome sequencing analysis, as well as Dr Caroline Schluth-Bolard and Dr Nicolas Chartron for the iPSC karyotypes. We also thank Isabelle Grosjean from the iPS_PGNM platform for her help and advice in cultivating induced pluripotent stem cells, Bruno Estebe (Imagine Institute, Paris) for his help in the establishment of the CRISPR-modified iPSC, and Leonardo Beccari for the useful insight into organoid cultures. We thank also all GenDev team members for constructive discussions as well as several intern students who helped to optimize experiments and acquire preliminary data: Lisa Malaisé, Grégoire Colomer, Aurora Devillaz, Lily-May Droulers, Noémie Gilibert, and Wassim Ouchetto. Finally, we thank the GenCiTy platform from the CRNL, and especially Sandrine Blondel who helped with the confocal settings for image acquisition, and Anne Ruiz who helped with the flow cytometry setting and acquisition.

## Funding

This work was supported by CNRS, Inserm and Université Lyon 1 through recurrent funding; the Agence Nationale de la Recherche (no. ANR-18CE12-0007; no. ANR-22CE12-0007); the Fondation Jérôme Lejeune and the Fondation pour la recherche sur le Cerveau « Espoir en tête » (confocal microscope). J.G. was supported by the Ministère de l’Enseignement Supérieur et de la Recherche and by the Fondation pour la Recherche Médicale. T-Y C. was supported by a postdoctoral fellowship from Academia Sinica, Taiwan and T.K.T by the National Science and Technology Council (NSTC 112-2326-B001-010) and Academia Sinica (AS-IA-109-L04), Taiwan. E.B. was supported by an EMBO long-term fellowship (ALTF-284-2019), and V.H. by the Swiss National Foundation (SNSF) 310030_205087. S.T. was supported by Agence Nationale de la Recherche (no. ANR-17-CE16-0003-01).

## Competing interests

The authors report no competing interests.

## Supplementary material

Supplementary material is available online.

## Notes

### Competing Interest Statement

The authors have declared no competing interest.

### Funding Statement

This work was supported by CNRS, Inserm and Universite Lyon 1 through recurrent funding; the Agence Nationale de la Recherche (no. ANR-18CE12-0007; no. ANR-22CE12-0007); the Fondation Jerome Lejeune and the Fondation pour la recherche sur le Cerveau "Espoir en tete" (confocal microscope). J.G. was supported by the Ministere de l'Enseignement Superieur et de la Recherche and by the Fondation pour la Recherche Medicale. T-Y C. was supported by a postdoctoral fellowship from Academia Sinica, Taiwan and T.K.T by the National Science and Technology Council (NSTC 112-2326-B001-010) and Academia Sinica (AS-IA-109-L04), Taiwan. E.B. was supported by an EMBO long-term fellowship (ALTF-284-2019), and V.H. by the Swiss National Foundation (SNSF) 310030_205087. S.T. was supported by Agence Nationale de la Recherche (no. ANR-17-CE16-0003-01).

### Author Declarations

French national ethical committee Comite de Protection des Personnes (number 2021-A01551-40) gave ethical approval for this work.

