## Supplementary data for "A TALS-like *RTTN* mutation impedes neural rosette formation in human cortical organoids"

**This PDF file includes:**

Supplementary Methods

Supplementary Figures 1 to 9 and legends

Supplementary Tables 1 to 4

SI References

### Supplementary Methods

#### Genetic studies

Informed consent for genetic analysis was obtained from the couple. DNA was extracted from blood samples. Trio Exome Sequencing was performed and analysed at the University Hospital of Lyon. Library preparation was performed with the Medexome kit (Roche) following manufacturers' instructions, and paired-end 2x150 sequencing on a NextSeq500 instrument (Illumina). Genomic alignment against the hg19/GRCh37 assembly and variant calling were, respectively, done with BWA-MEM v.0.7.12 (Li and Durbin, 2009) and GATK HaplotypeCaller v.3.4 (Broad Institute, Boston, MA, USA) while QC were evaluated using DeCovA.[1] Only highly confident variants were kept for analysis (total depth >9; alternative allele depth >4; no strand bias; mosaicism >10%). Rare variants were defined by a minor allele frequency below 1% in gnomAD database. Data are available on request from the authors. A Sanger validation was performed to confirm the candidate variant and parental segregation (Supplementary Table 1).

#### RNA extraction, RT-PCR and RT-qPCR

RNA was either extracted with the Nucleospin Tissue kit (Macherey-Nagel, 740952.50) following manufacturer's instructions, or with chloroform-trizol standard protocol, and 1.5 µg of RNA was treated with 1 U/µg of DNase I (ThermoFisher, EN0521) for 10 minutes at 37°C. To stop the reaction, 1x of EDTA (ThermoFisher, EN0521) was added and incubated for 10 minutes at 60°C. Then, 1 µg of DNA-clear RNA was reverse transcribed using the GoScript kit (Promega, A5001) following manufacturer's instructions, with a mixture of oligodT and random primers. For PCR, 100 ng of cDNA was mixed with 1X GoTaq green master mix (Promega, M7122) and 0.5 µM primer mix (forward and reverse, Supplementary Table 1) in a total volume of 20 µL. For RT-qPCR, 6.25 or 25 ng of cDNA was used per reaction with 0.5 µM of primer mix (Supplementary Table 1) and 1X

ONEgreen fast qPCR premix (Ozyme, OZYA008) in a total volume of 20 µL, using the Rotor-gene Q (Qiagen).

#### Generation of RPE1-based doxycycline-inducible cell lines

To generate doxycycline-inducible expression of mutated RTTN-GFP in *RTTN*-dKO RPE1 cells, we used a previously reported construct consisting of the GFP-tagged *RTTN* cDNA cloned into pLVX-Tight-Puro vector.[2] The missense mutation p.Arg985Gly (leading to a protein isoform named RG) was generated by site-directed mutagenesis using the QuikChange kit, and the exon 23 deletion, causing the loss of amino-acids 962-985 (Δ23 protein isoform), was obtained by RT-PCR amplification from pFlag-tagged WT RTTN construct and sub-cloned into in-frame pLVX-Tight-Puro vector. Both constructs were confirmed by Sanger sequencing. Then, lentiviruses were produced as previously described,[2] and used to infect *RTTN*-dKO RPE1 Tet-On cells stably expressing rtTA. The infected cells were selected with puromycin (10 µg/ml) for 7 days or by cell sorter on the basis of GFP fluorescence and expanded. The expression of mutated RTTN-GFP (RG or Δ23) was induced by doxycycline (1 µg/ml) treatment as previously described[2] and verified by immunofluorescence and western blot (Supplementary Fig. 3A, B).

#### CRISPR/Cas9-mediated genome editing in iPS cells

For CRISPR/Cas9-mediated insertion of c.2953G>A variant in *RTTN* gene, cr-RNAs were designed using CRISPOR-tefor (http://crispor.tefor.net/) tool.[3] The guide RNA assembly was prepared as followed : 400 µM of each of the tracr-RNA (IDT, 1075928) and cr-RNA (IDT, Supplementary Table 1) were mixed and incubated in a thermocycler using the following program: 94°C for 4 min, 75 cycles of 94°C 5 sec + 93,5°C 5 sec with each cycle an increment of 1°C, 20°C for 7 min. For transfection using the Nucleofector 4D (Lonza), 400,000 single CAU2 iPS cells, previously incubated for 1h with 10 µM Y-27632, were mixed with the P3 primary cell solution and Supplement 1 (both from Lonza, V4XP-3032), 62 pmol HiFi Caspase9 (IDT, 1081060), 400 pmol guide RNA and 300 pmol ssODN (IDT, Supplementary Table 1). This ssODN matrix contains, in addition to the c.2953G>A variant, a nucleotide change at the PAM sequence, predicted to not alter *RTTN* expression nor splicing, which avoids further cutting from the Cas9. After the electric shock, cells were allowed to recover in hot mTesR™ Plus medium supplemented with 10 µM Y-27-632 for 10 minutes, before being transferred into laminin 521-coated wells (STEMCELL Technologies, 77004). Medium was changed every other day, without adding Y-27632, until cells reached confluence and transferred onto vitronectin-coated dish at very low density (12.5 or 25 cells/cm²) in mTesR™ Plus medium supplemented with 10% CloneR (STEMCELL Technologies, 05888). When colonies were large enough, each colony was scrapped to perform Sanger sequencing (Supplementary Fig. 5A). Sequencing showed that 20% of all screened iPSC colonies presented a bi-allelic knock-in (KI) of the variant. Four clones with normal morphology and the desired genotypes (two WT and two KI) were controlled for genomic integrity using iCSdigital probes from Stem Genomics (Supplementary Fig. 5B), sequencing of predicted off-targets by CRISPOR-tefor[3] (Supplementary Fig. 5C), and G-banding karyotyping (Supplementary Fig. 5D). All correctly expressed *RTTN* with the presence of the three different isoforms in both KI clones (Supplementary Fig. 5E-G).

#### Ultrastructural expansion microscopy (U-ExM)

The U-ExM protocol was performed as previously reported.[4,5] Briefly, cells were seeded at a density of 75,000 cells/cm² on coverslips and the following day, were incubated with molecular anchors (1.4% formaldehyde (Sigma, F8776) and 2% acrylamide (Sigma, A4058)) for 3h at 37°C. Coverslips were then placed on top of 38 µL of ice-cold monomeric solution (1X PBS, 23% sodium acrylate (AK Scientific, R426), 10% acrylamide, 0.1% N,N′-methylenbisacrylamide (Sigma, M1533)) with 5% TEMED (ThermoFisher, 17919) and 5% ammonium persulfate (APS, ThermoFisher, 17874) placed on Parafilm, and incubated for 1h at 37°C for polymerization to happen, followed by a denaturation at 95°C for 1h30. The following day, a quarter of gel was incubated with primary antibodies diluted in 1X PBS, 2% BSA and incubated for 3h at 37°C with orbital agitation. After three washes with 1X PBS + 0.1% Tween20, secondary antibodies were incubated as previously done for primary antibodies but in the dark. After three more washes, the gel was expanded in water overnight. Upon imaging, a small piece of gel was cut and put on top of a poly-D-lysine (0.1 mg/ml, A3890401, Gibco)-coated coverslip. The Zeiss LSM 800 confocal microscope was used to image the NSC while an inverted Leica Thunder microscope was used to image the fibroblasts.

### Supplementary figures and legends

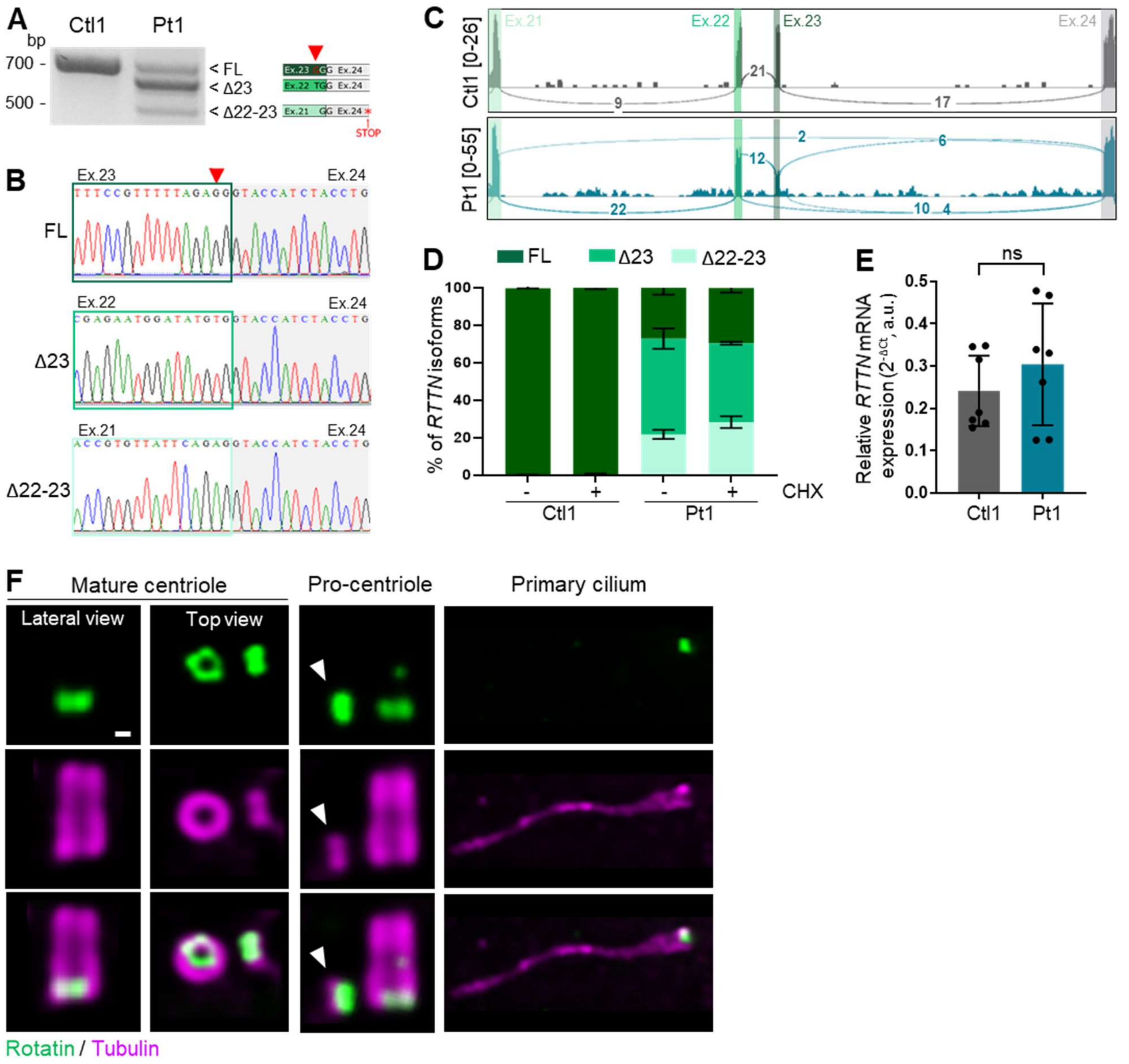


**Supplementary Figure 1. Characterization of the impact of c.2953A>G *RTTN* variant on *RTTN* pre-mRNA splicing and global expression in patient fibroblasts.** All experiments were performed in control (Ctl1) and patient (Pt1) fibroblasts. **(A)** RT-PCR analysis of exon 21 to exon 25 fragment in fibroblasts. Three splicing events are observed: a full-length (FL) form that includes the nucleotide change (red arrowhead), a form depleted of exon 23 (Δ23) and a form depleted of both exons 22 and 23 (Δ22-23) that leads to premature stop codons from exon 24. **(B)** Sanger sequencing of the patient PCR amplicons seen in A. **(C)** Sashimi plot visualization of aligned RNA sequencing data from fibroblasts at *RTTN* introns 21 to 23. On the arcs, the number of junction-spanning reads supporting exon pairs; in brackets, the range of coverage of each base of the depicted region**. (D-E)** RT-qPCR analyses of each of the *RTTN* splicing isoforms (D), in absence (-) or presence (+) of the NMD inhibitor cycloheximide (CHX), and of the relative global expression (E) in fibroblasts. *RPS17* was used as the house-keeping gene. Graph (E) shows the mean ± SD of seven independent experiments. Differences are not significant (ns) by Mann-Whitney's test. **(F)** Representative confocal images of Rotatin (green) localisation at expanded centrioles (lateral or top views) and at the base of primary cilium (α/β-Tubulin, magenta) in control fibroblasts. Scale bar: 100 nm. a.u., arbitrary units.

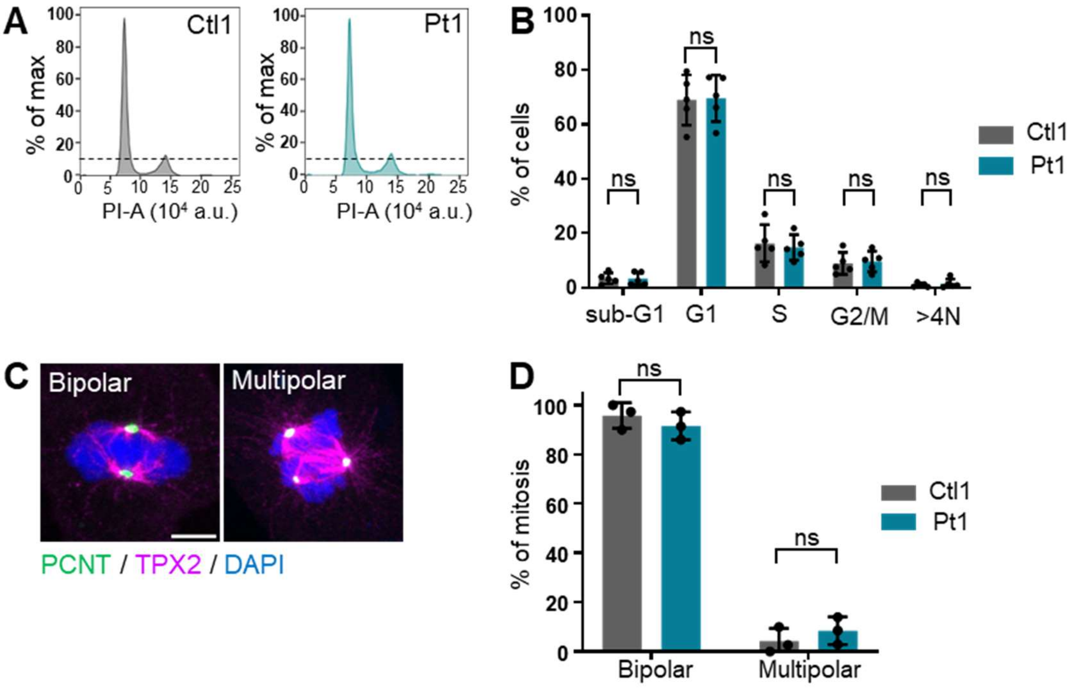


**Supplementary Figure 2. Characterization of the impact of c.2953A>G *RTTN* variant on cell cycle progression and mitosis in patient fibroblasts.** All experiments were performed in control (Ctl1) and patient (Pt1) fibroblasts. **(A)** Flow cytometric cell cycle analysis histograms in fibroblasts. The dotted line represents the top of G2/M peak in Ctl1 cells for reference. **(B)** Quantification of the percentage of cells in each cell cycle phase. Graph shows the mean ± SD of five independent experiments. **(C)** Confocal images of representative normal (bipolar) and abnormal (multipolar) mitosis events seen in fibroblasts. Pericentrin (PCNT) stains centrosomes, TPX2 mitotic spindles and DAPI DNA. **(D)** Quantification of the proportion of normal and abnormal mitosis events such as seen in C. Graph shows the mean ± SD of three independent experiments (n>100 mitosis per experiment). Differences are not significant (ns) by two-way ANOVA with Tukey's correction (B, D). Scale bar: 5 µm. a.u., arbitrary units; PI, propidium iodide.

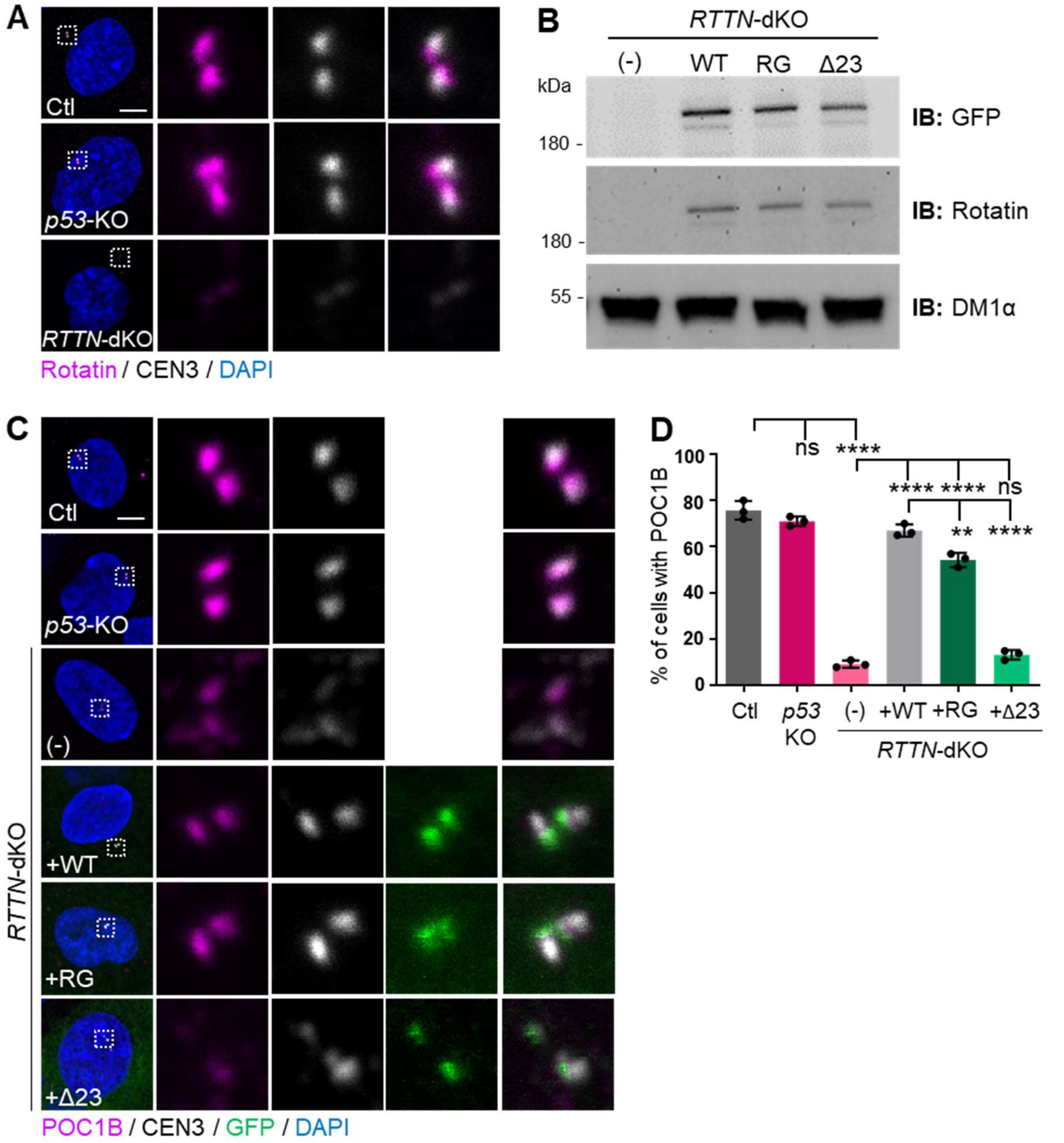


**Supplementary Figure 3. Characterization of *RTTN*-dKO RPE1 cellular model and analysis of POC1B centriolar localisation.** All experiments were performed in control, *p53*-KO and *RTTN*-dKO RPE1 cells induced to express wild-type (WT) or mutated (RG, Δ23) RTTN-GFP proteins. **(A)** Representative confocal images of Rotatin protein expression (magenta) in *RTTN*-dKO RPE1 cellular model. CEN3 labels centrioles. **(B)** Western blot analysis of Rotatin protein expression, with the indicated antbodies, in *RTTN*-dKO RPE1 cellular model. α-Tubulin (DM1α) is used as a loading control. **(C)** Confocal images of POC1B (magenta) localization at centrosomes (CEN3, grey) in *RTTN*-dKO RPE1 cellular model. **(D)** Quantitative analysis of cells expressing POC1B protein at the centrioles such as seen in C. Graph shows the mean ± SD from three independent experiments (100 cells per experiment). ns, not significant; **p-value< 0.01; ****p-value<0.0001 following one-way ANOVA with Tukey's multiple comparison test. Scale bar: 5 µm. DAPI stains DNA.

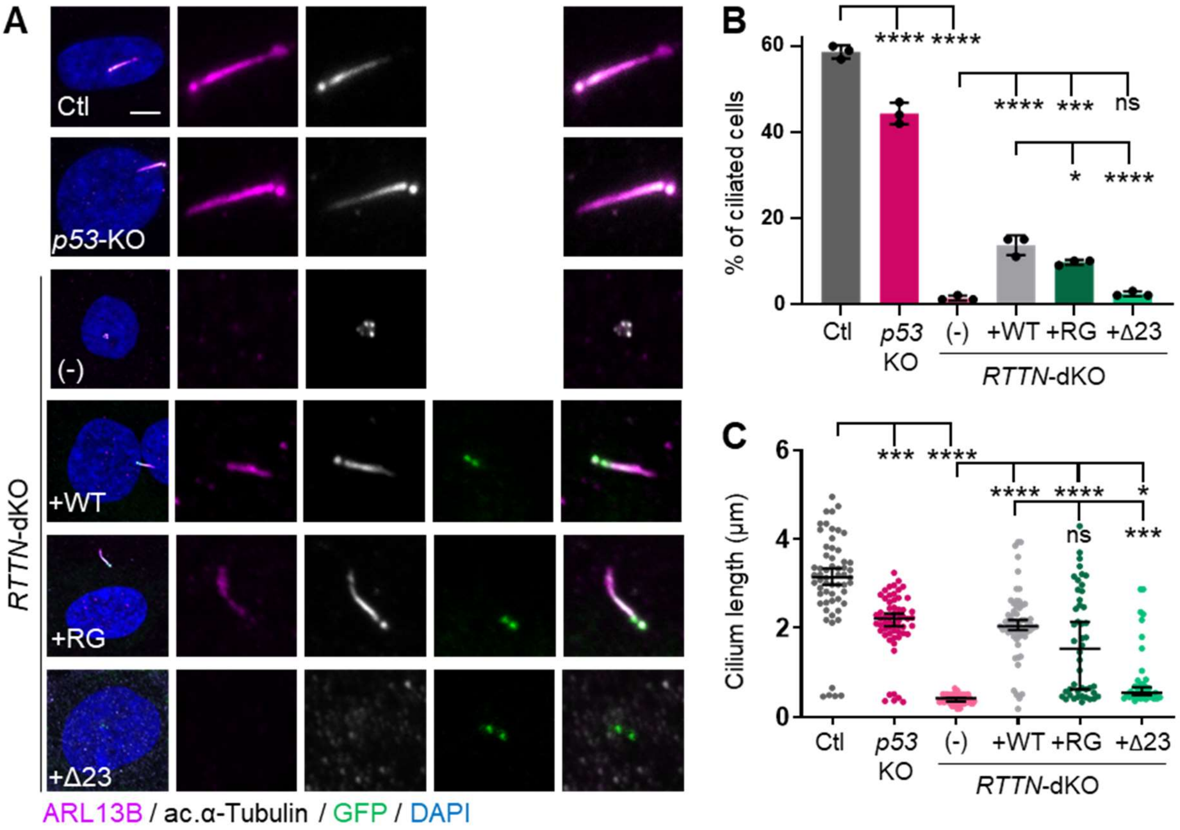


**Supplementary Figure 4. Characterization of the impact of p.Arg985Gly (RG) and Δ23 variants on primary cilium formation in *RTTN*-dKO RPE1 cellular model.** All experiments were performed in control, *p53*-KO and *RTTN*-dKO RPE1 cells induced to express wild-type (WT) or mutated (RG, Δ23) RTTN-GFP proteins. **(A)** Representative confocal images of primary cilium, stained with the ciliary membrane marker ARL13B (magenta) and the axonemal marker acetylated-αTubulin (grey), in *RTTN*-dKO RPE1 cellular model. DAPI (blue) labels DNA. **(B-C)** Quantification of the percentage of ciliated cells (B) and length of primary cilium (C) such as seen in A. Graphs show the mean ± SD (B) or the median ± 95% CI (C) of three independent experiments (n>40 cells). ns, not significant; *p-value<0.05; ***p-value<0.005; ****p-value<0.0001 following one-way ANOVA with Dunnett's multiple comparison test (B) or Kruskal-Wallis with Dunn's multiple comparison test (C). Scale bar: 5 μm.

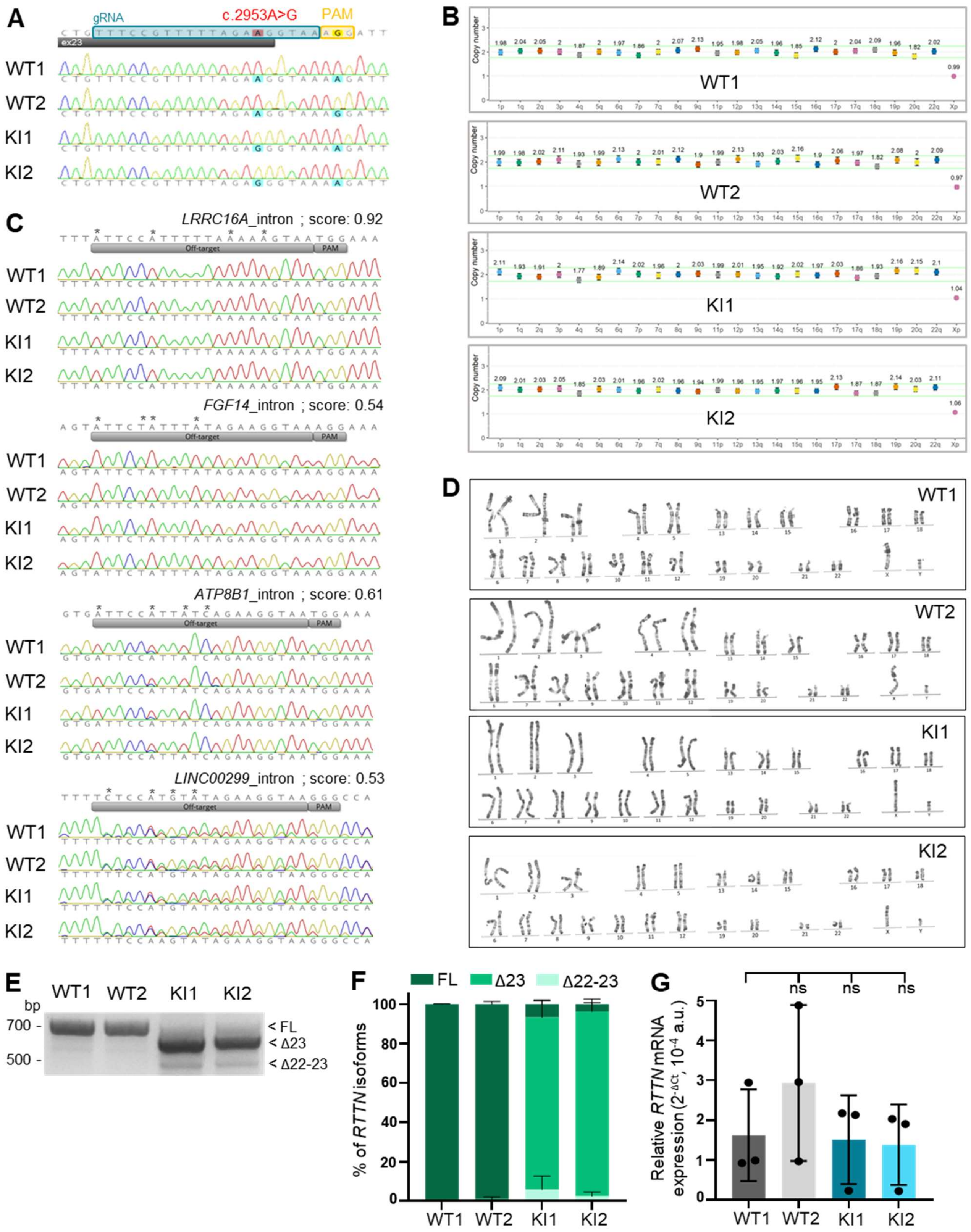


**Supplementary Figure 5. Validation of the CRISPR/Cas9-mediated genetic modification of *RTTN* gene in four iPS clones.** All experiments were performed in two wild-type (WT) and two knock-in (KI) iPS clones. **(A)** Chromatograms of Sanger sequencing of iPS clones showing the region encompassing the *RTTN* c.2953A nucleotide. Positions of the guide RNA and the PAM sequences are highlighted in blue and yellow, respectively. **(B)** Analysis of the copy number of the most frequent chromosomic variations seen in iPS clones, using iCS-digital™ PCS test. Each marker is seen in two copies, except the marker on chr. X (male genotype), validating the genomic integrity of the iPS clones. **(C)** Chromatograms of Sanger sequencing of the off-targets predicted with the highest score (>0.4). Asterisks highlight the nucleotide mismatches between the *RTTN* targeted sequence and the off-target. No mutation in the introns of the tested genes was detected. **(D)** G-banding karyotypes of the selected iPS clones. No chromosomic alteration is observed. **(E-F)** RT-PCR (E) and RT-qPCR (F) analyses of the splicing events of *RTTN* exon 23 in iPS clones. **(G)** RT-qPCR analysis of *RTTN* relative expression in iPSC clones normalised to *RPS17*. Graphs (F, G) show the mean ± SD of three independent experiments. Differences are not significant by Kruskal-Wallis test with Dunn's multiple comparisons test. a.u. arbitrary units.

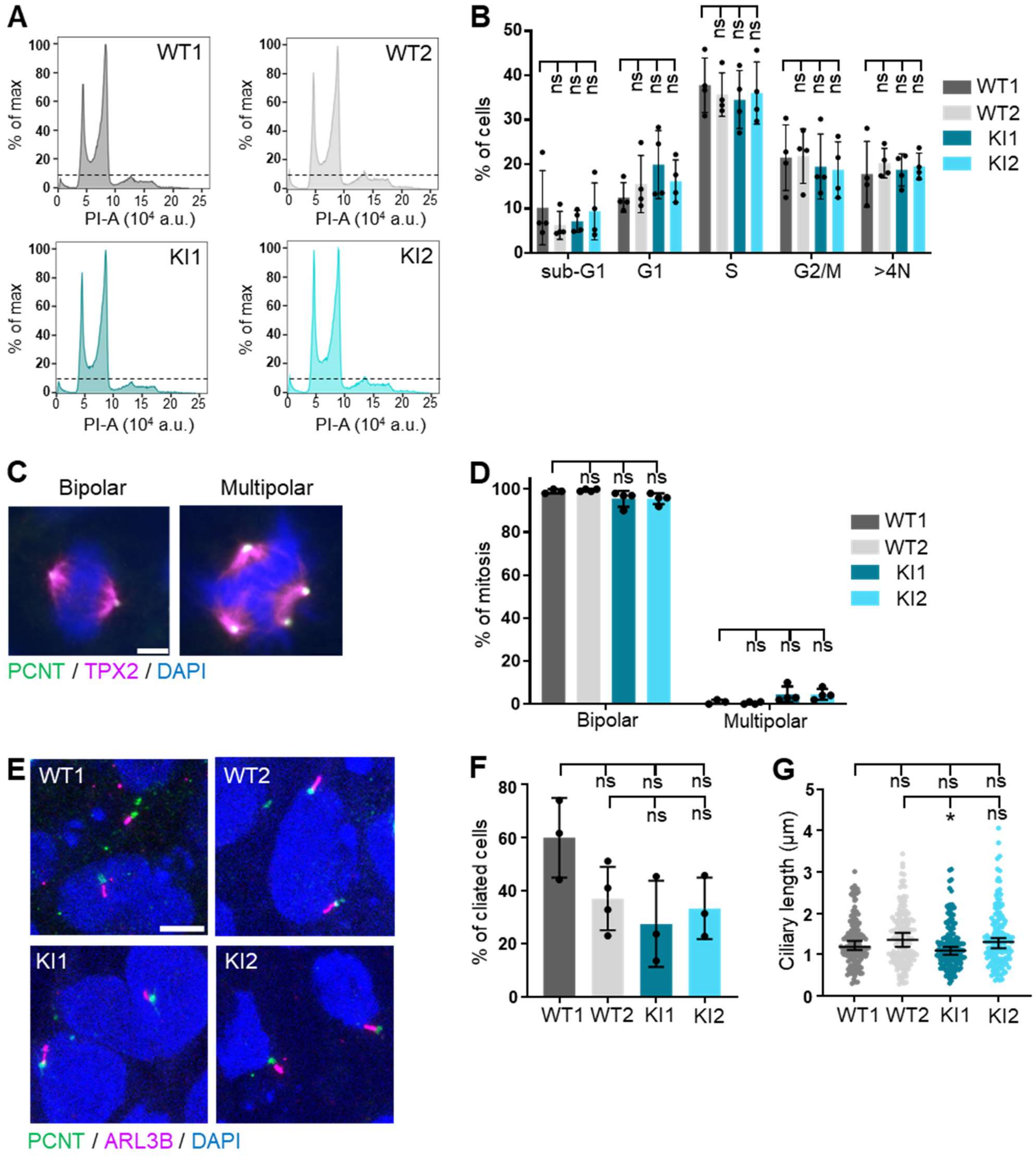


**Supplementary Figure 6. Characterization of *RTTN*-mutated iPS clone phenotypes.** All experiments were performed in two wild-type (WT) and two knock-in (KI) iPS clones. **(A)** Flow cytometric cell cycle analysis histograms in iPS clones. **(B)** Quantification of the percentage of cells in each cell cycle phase. Graph shows the mean ± SD of four independent experiments. **(C)** Confocal images of representative normal (bipolar) and abnormal (multipolar) mitosis events seen in iPS clones. Pericentrin (PCNT) stains centrosomes, TPX2 mitotic spindles. **(D)** Quantification of the proportion of normal and abnormal mitosis events such as seen in C. Graph shows the mean ± SD of four independent experiments (n>100 mitosis per experiment). **(E)** Representative confocal images of primary cilium (ARL13B, magenta) and centrosome (PCNT, green) in iPS clones. **(F, G)** Quantification of the percentage of ciliated cells (F) and of length of primary cilium (G) such as observed in E. Graphs show the mean ± SD (F) or median ± 95% CI (G) of four independent experiments (n=150 cells per experiment). ns, non-significant; *p-value<0.5 by two-way (B, D) or one-way (F) ANOVA with Tukey's correction, or Kruskal-Wallis test with Dunn's multiple comparisons test (G). Scale bars: 2 µm (B), 5 µm (C). DAPI labels DNA. a.u. arbitrary units.

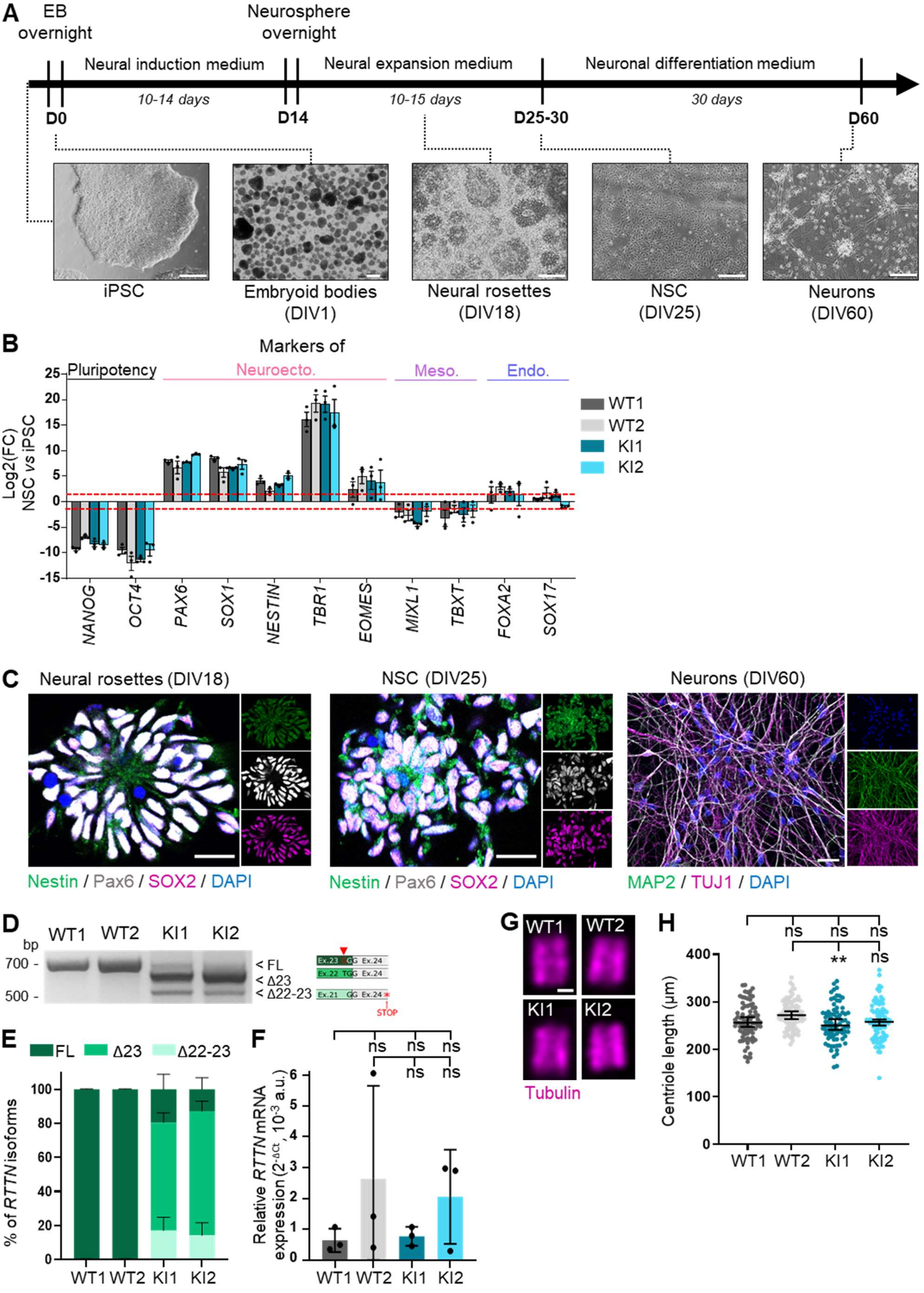
A
EB overnight
Neurosphere overnight
Neural induction medium
Neural expansion medium
Neuronal differentiation medium
10-14 days
10-15 days
30 days
D0
D14
D25-30
D60
iPSC
Embryoid bodies (DIV1)
Neural rosettes (DIV18)
NSC (DIV25)
Neurons (DIV60)
B
Markers of
Pluripotency
Neuroecto.
Meso.
Endo.
Log2(FC) NSC vs iPSC
WT1
WT2
KI1
KI2
NANOG
OCT4
PAX6
SOX1
NESTIN
TBR1
EOMES
MIXL1
TBXT
FOXA2
SOX17
C
Neural rosettes (DIV18)
NSC (DIV25)
Neurons (DIV60)
Nestin / Pax6 / SOX2 / DAPI
Nestin / Pax6 / SOX2 / DAPI
MAP2 / TUJ1 / DAPI
D
WT1 WT2 KI1 KI2
bp
700 -
500 -
< FL
< Δ23
< Δ22-23
STOP
E
FL Δ23 Δ22-23
% of RTTN isoforms
WT1 WT2 KI1 KI2
F
Relative RTTN mRNA expression (2-ΔCt, 10-3 a.u.)
ns ns ns
ns ns
WT1 WT2 KI1 KI2
G
WT1 WT2
KI1 KI2
Tubulin
H
ns ns ns
** ns
Centriole length (μm)
WT1 WT2 KI1 KI2

**Supplementary Figure 7. 2D differentiation of iPSC into neural stem cells (NSC) and neurons.** All experiments were performed in control (WT) and *RTTN*-mutated (KI) NSC. **(A)** Timeline of iPSC differentiation into neural stem cells and neurons. Bright field images of cells at the key stages of the differentiation are shown. **(B)** RT-qPCR analysis of gene expression of markers of pluripotency, neuroectoderm, mesoderm and endoderm in NSC compared to iPSC. iPSC express markers of pluripotency while NSC predominantly express markers of neuroectoderm. Graph shows the mean ± SD of three independent experiments. **(C)** Representative confocal images of neural rosettes (left), NSC (middle) and neurons (right). Neural rosettes and NSC are stained for nestin (green), Pax6 (grey) and SOX2 (magenta) while neurons are labelled with MAP2 (green) and TUJ1 (magenta). DAPI stains nuclei. **(D-F)** RT-PCR (D) and RT-qPCR (E) analyses of the splicing events of *RTTN* exon 23, and of the *RTTN* relative expression in NSC. *RPS17* was used as a house-keeping gene. Graphs show the mean ± SD of three independent experiments. **(G)** Representative confocal images of expanded centrioles (Tubulin, magenta) in NSC. **(H)** Quantification of length of centriole such as seen in G. Graph shows the median ± 95% CI of three independent experiments. ns not significant; **p-value<0.01 following Kruskal-Wallis test with Dunn's multiple comparisons test (F) or one-way ANOVA with Tukey's correction (H). Scale bars: 200 µm (A), 20 µm (C), 100 nm (G). a.u. arbitrary units; EB, embryoid bodies.

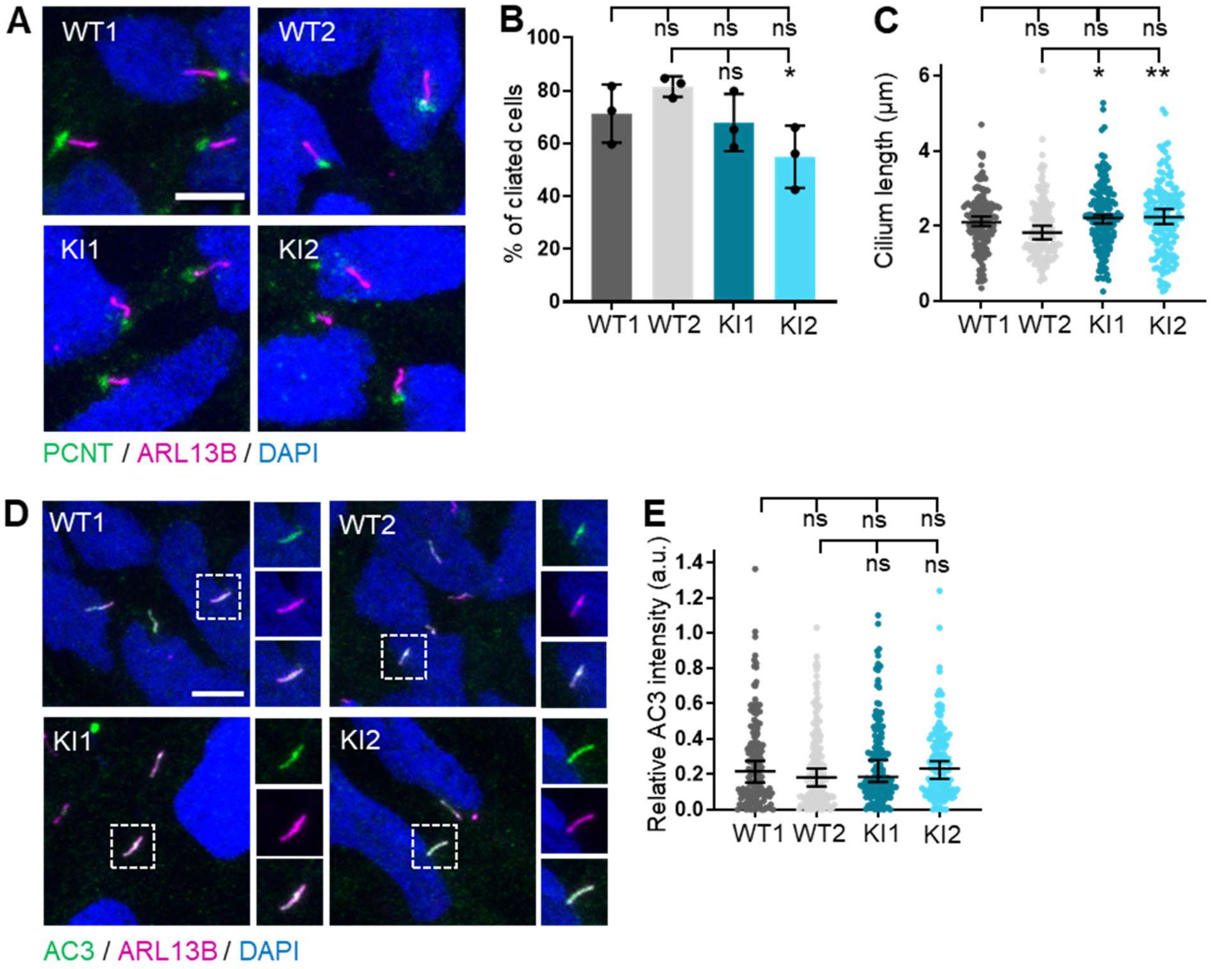


**Supplementary Figure 8. Characterization of primary cilium formation and function in *RTTN*-mutated NSC.** All experiments were performed in control (WT) and *RTTN*-mutated (KI) NSC. **(A)** Representative confocal images of primary cilium (ARL13B, magenta) and centrosome (PCNT, green) in NSC. **(B, C)** Quantification of percentage of ciliated cells (B) and of length of primary cilium (C) such as seen in A. Graphs show the mean ± SD (B) or median ± 95% CI (C) of three independent experiments (n=150 cells). **(D)** Representative confocal images of adenylate cyclase 3 (AC3, green) in primary cilium (ARL13B, magenta) in NSC. **(E)** Quantification of relative AC3 intensity compared to ARL13B. Graph shows the median ± 95% CI of three independent experiments (n=150 cells). ns not significant; *p-value<0.05; **p-value<0.01 following one-way ANOVA with Tukey's correction (B) or Kruskal-Wallis with Dunn's correction (C, E). Scale bars: 5µm. DAPI labels the nuclei. a.u., arbitrary units.

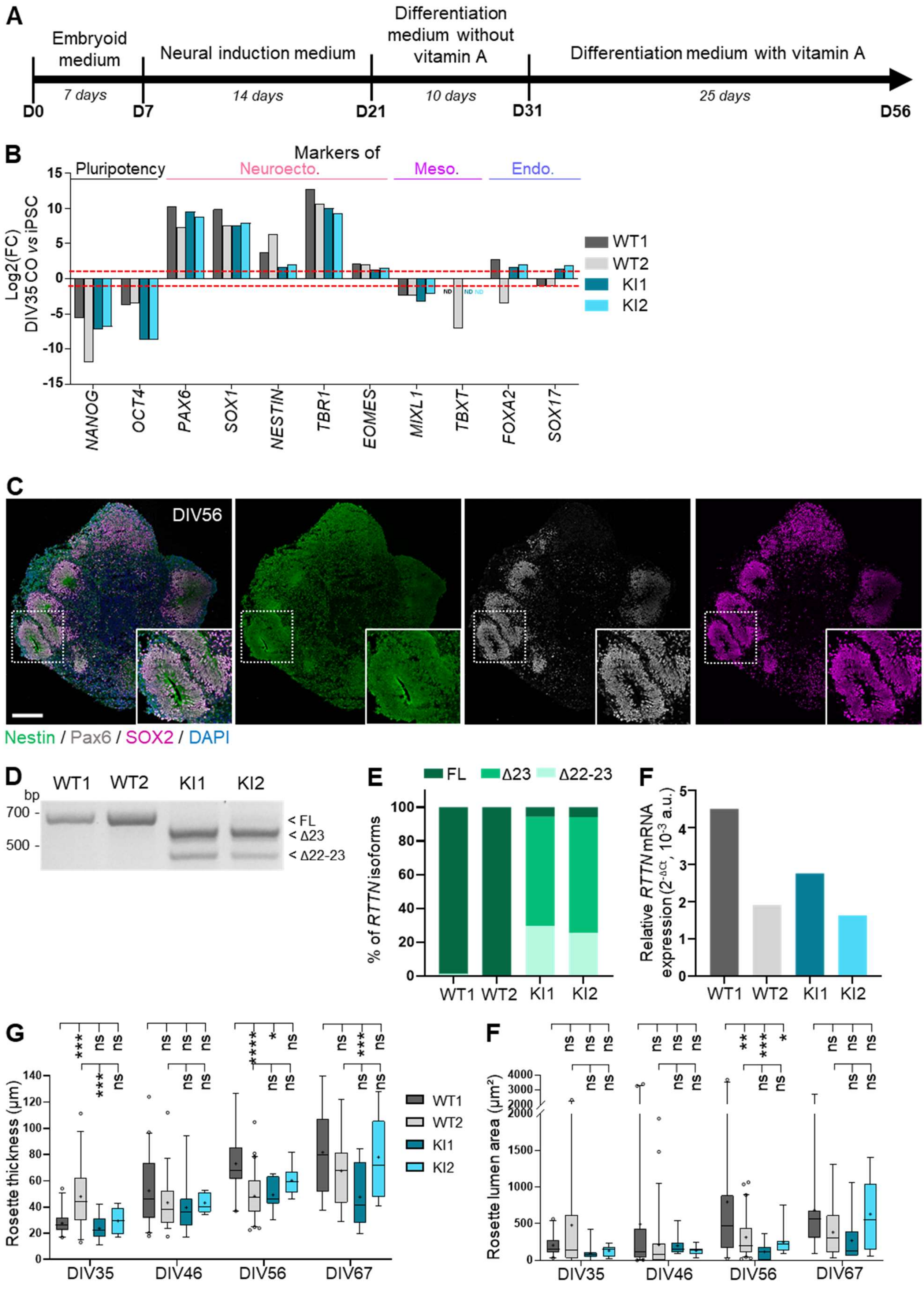


**Supplementary Figure 9: 3D differentiation of iPSC into cortical organoids.** All experiments were performed in wild-type (WT) and *RTTN*-mutated (KI) cortical organoids (CO). **(A)** Timeline of iPSC differentiation into cortical organoids. **(B)** RT-qPCR analysis of gene expression of markers of pluripotency, neuroectoderm, mesoderm and endoderm in CO at DIV35 compared to iPSC. iPSC express markers of pluripotency while CO predominantly express markers of neuroectoderm. Graph shows the results of one single experiment. **(C)** Representative confocal images of wild-type CO at DIV56. Nestin (green), Pax6 (grey) and SOX2 (magenta) label the NSC organized into neural rosettes (insets). DAPI stains nuclei. **(D-F)** RT-PCR (D) and RT-qPCR (E) analyses of the splicing events of *RTTN* exon 23, and of *RTTN* relative expression in CO at DIV35. *RPS17* was used as a house-keeping gene. Graphs show one single experiment of the pool of 8 organoids. **(G, H)** Quantification of thickness (G) and lumen area (H) of rosettes such as seen in Figure 6A. Box-and-whisker plots show in the box the median (the mean by the cross) and the 25th-75th percentiles, and in whiskers the 5th to 95th percentile of values from 4 different organoids (n=6 to 20 slices total). ns, non-significant; *p-value<0.05; **p-value<0.01; ***p-value<0.001; ***p-value<0.0001 following two-way ANOVA with Tukey's correction (G, F). Scale bar: 200 µm. a.u., arbitrary units; ND, not detected.

**Supplementary Table 1. Primer sequences**

| | | |
|---|---|---|
| | **RT-PCR primer sequences for *RTTN*** | |
| **Targeted region** | **Primer F** | **Primer R** |
| Exon 21 – Exon 25 | TGTGTGAGTCAAGATGGCAAG | TTGCAGCCTGAACAATGGAATG |
| | **RT-qPCR primer sequences for *RTTN*** | |
| **Targeted isoform(s)** | **Primer F** | **Primer R** |
| All | CATCAGCATTTGTTCAAAGATCTG | GCAGTGGTAAGTTTCAGCTTTCA |
| Full-length | CTGTTTCCGTTTTTAGAAGGTAC | |
| Δ23 | ATCGAGAATGGATATGTGGTAC | CGGCTTCAAGGCCAAACAAT[a] |
| Δ22-23 | CCGTGTTATTCAGAGGTACCA | |
| | **PCR primer sequences to sequence gDNA after CRISPR-Cas9 experiment** | |
| **Gene** | **Primer F** | **Primer R** |
| *RTTN* | TCATTTGCAGTCAACGAAGTGAG | AGGACCATGAAAGCCCAAGTTA |
| *FGF14* | CATCCTTGGTTCCATCGAGA | CCCTGAGCTCACAAGTAGAG |
| *LRRC16A* | ATACAACACCTGGCGTTAGGC | CAGGAGCAACGAAAGGGGATTA |
| *ATP8B1* | CACTAGAAGCTATAAACGCACTTC | TTAGTCCAAGTGTCTTCCAGCAG |
| *LINC00299* | ATGAGCCCTTTTCCTAGATCTGAC | CTCAGAAAAGGGAGCTAAGAAGC |
| | **RT-qPCR primer sequences to analyze expression of differentiation markers** | |
| **Gene** | **Primer F** | **Primer R** |
| *RPS17* | CATTATCCCCAGCAAAAAGC | AGGCTGAGACCTCAGGAACA |
| *NANOG* | AAATACCTCAGCCTCCAGCAG | TGCGTCACACCATTGCTATTC |
| *OCT4* | AAACCCACACTGCAGCAGATCA | TCCTCTCGTTGTGCATAGTCG |
| *PAX6* | CCAACCAATTCCACAACCCA | GTGAGGGCTGTGTCTGTTCG |
| *SOX1* | GCTGACACCAGACTTGGGTTT | CCCCTCGAGCAAAGAAAACG |
| *EOMES* | GTGGCAAAGCCGACAATAACA | CCTGTCTCATCCAGTGGGAAC |
| *TBR1* | CGTGCAGACGTTCACTTTCC | TGTAATATCCGTGTTCTGGTAGGC |
| *NESTIN* | CTCAGCTTTCAGGACCCCAAG | GCAAAGATCCAAGACGCCG |
| *TBXT* | GCTTCAAGGAGCTCACCAATG | AGACACGTTCACCTTCAGCA |
| *MIXL1* | GCAAGCGCACGTCTTTCAG | CGCAAGTGGATGTCGGGGTA |
| *SOX17* | ATGTGTCCCAAAACAGCTTCC | ACACACCCAGGACAACATTTCT |
| *FOXA2* | CCCCCTACGCCAACATGAAC | TAGCTGCGCCTGTAGGTCTT |
| | **crRNA and ssODN sequences for *RTTN* editing by CRISPR-Cas9** | |
| crRNA[b] | 3’- TTTCCGTTTTTAGAAGGTAA *AGG* -5’ | |
| ssODN[c] | GTCTTCAGTTTGCCTGTTTCCGTTTTTAGAGGgtaaaagatttatttctctgattttcttgaaggacg | |

[a]Same reverse primer for all three transcripts

[b]PAM sequence in italic

[c]Underlined nucleotides are those modified to introduce the c.2953A>G variant and to alter the PAM

**Supplementary Table 2. Culture media for 2D neuronal differentiation protocol**

| Product | Supplier | Catalog number | Initial concentration | Final concentration | Dilution |
|---|---|---|---|---|---|
| **N2B27 Medium** | | | | | |
| DMEM/F12 | Gibco | 21331020 | - | 50% | 1/2 |
| Neurobasal | Gibco | 21103049 | - | 50% | 1/2 |
| N2 | Gibco | 17402048 | 100X | 1X | 1/100 |
| B27 without vitamin A | Gibco | 12587010 | 50X | 1X | 1/50 |
| Penicilin-streptomycin | Gibco | 15140122 | - | 1% | 1/100 |
| L-glutamine | Gibco | 25030024 | 200mM | 2mM | 1/100 |
| **Neural Induction Medium (NIM)** | | | | | |
| N2B27 | - | - | - | 100% | 1 |
| β-mercaptoethanol | Gibco | 31350010 | 50mM | 100μM | 1/500 |
| FGF2 | STEMCELL Technologies | 78003.1 | 1μg/mL | 20ng/mL | 1/50 |
| LDN-193189 | STEMCELL Technologies | 72147 | 1mM | 500nM | 1/2000 |
| SB-431542 | STEMCELL Technologies | 72234 | 10mM | 20μM | 1/500 |
| **Neural Expansion Medium (NEM)** | | | | | |
| N2B27 | - | - | - | 100% | 1 |
| FGF2 | STEMCELL Technologies | 78003.1 | 1μg/mL | 20ng/mL | 1/50 |
| EGF | STEMCELL Technologies | 78006.1 | 10μg/mL | 10ng/mL | 1/1000 |
| BDNF | STEMCELL Technologies | 78005 | 10μg/mL | 20ng/mL | 1/500 |

FGF2, Fibroblast growth factor 2; EGF, Epidermal growth factor; BDNF, Brain derived neuronal factor

**Supplementary Table 3. Culture media for cortical organoids**

| Product | Supplier | Catalog number | Initial concentration | Final concentration | Dilution |
|---|---|---|---|---|---|
| **Embryoid Body Medium (EB)** | | | | | |
| mTeSR™ Plus | STEMCELL Technologies | 100-0276 | - | - | 1 |
| Y-27632 | STEMCELL Technologies | 72302 | 10mM | 10μM | 1/1000 |
| SB-431542 | STEMCELL Technologies | 72234 | 10mM | 10μM | 1/1000 |
| **Neural Induction Media (NIM)** | | | | | |
| DMEM-F12 | Gibco | 21331020 | - | - | 1 |
| L-glutamine | Gibco | 25030024 | 200mM | 2mM | 1/100 |
| N2 | Gibco | 17402048 | 100X | 1X | 1/100 |
| Non Essential Amino Acid | Gibco | 11140035 | 100X | 1X | 1/100 |
| Penicilin-streptomycin | Gibco | 15140122 | - | 0.1% | 1/1000 |
| β-mercaptoethanol | Gibco | 31350010 | 50mM | 100μM | 1/500 |
| LDN-193189 | STEMCELL Technologies | 72147 | 1mM | 250nM | 1/4000 |
| SB-431542 | STEMCELL Technologies | 72234 | 10mM | 10μM | 1/1000 |
| **Neural Expansion Media (NEM)** | | | | | |
| DMEM-F12 | Gibco | 21331020 | - | 50% | 1/2 |
| Neurobasal | Gibco | 21103049 | - | 50% | 1/2 |
| N2 | Gibco | 17402048 | 100X | 0.5X | 1/200 |
| B27, with vitamin A (DM1)[a] | Gibco | 17504044 | 50X | 0.5X | 1/100 |
| B27, without vitamin A (DM2)[a] | Gibco | 12587010 | 50X | 0.5X | 1/100 |
| L-glutamine | Gibco | 25030024 | 200mM | 2mM | 1/100 |
| Penicilin-streptomycin | Gibco | 15140122 | - | 0.1% | 1/1000 |
| Non Essential Amino Acid | Gibco | 11140035 | 100X | 1X | 1/100 |
| β-mercaptoethanol | Gibco | 31350010 | 50mM | 100μM | 1/500 |
| Insuline | Sigma Aldrich | I9278 | See lot | 6.25μg/mL | See lot |

[a]DM1 from DIV21 to DIV30; DM2 from DIV31 to DIV67

**Supplementary Table 4. Antibodies**

| Primary antibodies | | | | |
|---|---|---|---|---|
| **Targeted protein** | **Supplier** | **Catalog number** | **Host species** | **Dilution** |
| Adenylate cyclase 3 | Fisher Scientific | 15368114 | Rabbit | 1/500 |
| ARL13B | Proteintech | 17711-1 AP | Rabbit | 1/500 |
| Cleaved caspase 3 | Cell signaling | 9661 | Rabbit | 1/500 |
| Centrin3 | Abnova | H00001070-M01 | Mouse | 1/5,000 |
| DM1 α-tubulin | Sigma Aldrich | T9026 | Mouse | 1/10,000 |
| GFP | BD Biosciences | 632381 | Mouse | 1/5,000 |
| Ki67 | BD Biosciences | 550609 | Mouse | 1/250 |
| MAP2 | Abcam | ab5392 | Chicken | 1/1000 |
| N-cadherin | BD Biosciences | 610921 | Mouse | 1/500 |
| Nestin | Novus/Bio-techne | NB100-1604 | Chicken | 1/500 |
| p21 | Fisher Scientific | R.229.6 | Rabbit | 1/500 |
| p53 | Fisher Scientific | DO-7 | Mouse | 1/200 |
| Pax6 | BD Biosciences | 561462 | Mouse | 1/100 |
| Pericentrin | Abcam | ab220784 | Rabbit | 1/750 |
| Phospho-vimentin | Clinisciences | D076-3 | Mouse | 1/500 |
| POC1B | Thermofisher | PA5-24495 | Rabbit | 1/400 |
| Rotatin | Tang's lab - Taiwan | PMID: 28811500 | Rabbit | 1/400 |
| Sox2 | Invitrogen | 14-9811-82 | Rat | 1/250 |
| TPX2 | Bio-techne | NB500-179 | Rabbit | 1/500 |
| Acetylated α-tubulin | Sigma Aldrich | T6793 | Mouse | 1/1,000 ; 1/125 (UExM) |
| Acetylated α-tubulin | Sigma Aldrich | T7451 | Mouse | 1/400 |
| TUJ1 | Thermofisher | 15234347 | Mouse | 1/1,000 |
| ZO-1 | BD Biosciences | 610966 | Mouse | 1/300 |

| Secondary antibodies | | | | |
|---|---|---|---|---|
| **Targeted species** | **Fluorophore** | **Supplier** | **Catalog number** | **Dilution** |
| Goat anti-chicken | Alexa 488 | Invitrogen | A11039 | 1/1,000 |
| Goat anti-rabbit | Alexa 488+ | Invitrogen | A32731 | 1/1,000 |
| Goat anti-mouse | Alexa 488+ | ThermoFisher | A32723 | 1/1,000 |
| Goat anti-rabbit | Alexa 555+ | ThermoFisher | A32732 | 1/1,000 |
| Goat anti-mouse | Alexa 555+ | ThermoFisher | A32727 | 1/1,000 ; 1/400 (UExM) |
| Goat anti-rabbit | Alexa 647+ | ThermoFisher | A32733 | 1/1,000 |
| Goat anti-mouse | Alexa 647+ | Invitrogen | A32728 | 1/1,000 |
| Goat anti-rat | Alexa 647+ | Thermofisher | A21247 | 1/1,000 |
